# Dissecting the effect of long-term exposure to air pollution on risk of dementia in UK Biobank

**DOI:** 10.1101/2025.06.18.25329828

**Authors:** Ensor Rafael Palacios, Chin Yang Shapland, Levi John Wolf, Liv Tybjærg Nordestgaard, Emma Anderson, Chloe Slaney, Dan Bernie, Dann Mitchell, Patrick Gavin Kehoe, Gareth John Griffith, Kate Tilling

**Affiliations:** MRC Integrative Epidemiology Unit, University of Bristol, Beacon House, Bristol, BS8 1QU, UK; School of Geographical Sciences, University of Bristol, Beacon House, Bristol, BS8 1QU, UK; Department Of Clinical Biochemistry, Copenhagen University Hospital, Blegdamsvej 3B, Copenhagen, 2200, Denmark; Department of Mental Health of Older People, UCL, Gower Street, London, WC1E 6BT, UK; Met Office, Fitzroy Road, Exeter, EX1 3PB, UK; Faculty of Health Sciences, University of Bristol, Beacon House, Bristol, BS8 1QU, UK; Cabot Institute for the Environment, University of Bristol, Beacon House, Bristol, BS8 1QU, UK; Translational Health Sciences, University of Bristol, Beacon House, Bristol, BS8 1QU, UK

**Keywords:** air pollution, dementia, Alzheimer’s disease, vascular dementia, environmental epidemiology

## Abstract

Mounting evidence links air pollution to dementia, the most prevalent cause of cognitive impairment in older people. Here we investigated individual and compound effects of particulate matters (PM_10_, PM_2.5_, PM*_coarse_*, PM*_abs_*) and nitric oxides (NO_2_, NO) on risk of all-cause dementia, and its most common subtypes, Alzheimer’s disease (AD) and vascular dementia (VAD), using data from UK Biobank. We addressed factors that hinder causal interpretation of associations previously shown in the literature and their translation into clear public health policies. Specifically: 1) spatial confounding by area-level covariates, 2) collinearity among and identification of the most relevant air pollutants, and 3) the time window for pollution exposure. Furthermore, we used chronic obstructive pulmonary disease (COPD) and frequency of oily fish intake in positive and negative control analyses. We found NO_2_ to be the strongest risk factor for dementia, especially when considering longer periods (≥ 5 years) of exposure (all-cause dementia hazard ratio HR=1.06, 1.02-1.11 per 9.86 *µg/m*^3^ interquartile range). There was stronger evidence of an effect on risk for AD than VAD. Positive control analysis did not provide any evidence against causality, although the analyses of spatial confounding and negative control analyses revealed the presence of some residual bias, thus warranting care in the interpretation of the results. Together, our results highlight that targeting air pollution, in particular NO_2_ levels, could inform preventive public health policies for dementia.

## 1 Introduction

Dementia is a syndrome characterised by the progressive decline of cognitive, emotional, and motor abilities due to accruing brain damage. It eventually impairs a person’s everyday life to a point where extensive support is needed, thereby placing a huge burden on the caregivers and the health system [1]. Currently, there are over 50 million people suffering from dementia worldwide, a number that is predicted to triple by 2050 [1, 2], if the current absence of effective treatments and prevention strategies continues.

One proposed risk factor for dementia is air pollution, carrying a smaller risk for individuals but with higher population prevalence than other factors (e.g., diabetes, obesity, hearing loss) [3]. Air pollution encompasses chemicals, gases, and particles from various natural and anthropogenic sources, including particulate matter (PM) and nitrogen oxides (NO*_x_*) [4]. PMs are classified based on their composition (e.g., black carbon and soot, PM*_abs_*) and aerodynamic diameter (≤ 10*µm*, PM_10_, 2.5 − 10*µm*, PM*_coarse_*, ≤ 2.5*µm*, PM_2.5_), with smaller particles able to penetrate deeper into human tissues (e.g., by crossing the blood brain barrier) [5, 6]. NO*_x_* includes nitric oxide (NO) and dioxide (NO_2_), which are highly reactive species that interact primarily with the components of the respiratory system [5]. The main pathways through which air pollution can affect the brain involve increased oxidative stress and inflammatory responses, which trigger cascade effects leading to tissue damage [7–9]. Notably, air pollution may unequally affect dementia subtypes, including Alzheimer’s disease (AD) and vascular dementia (VAD), because their partially distinct underlying neurodegenerative mechanisms (protein structure vs neurovascular alterations) could be differentially susceptible to pollutants toxicity.

Growing epidemiological evidence supports an adverse effect of air pollution on risk of dementia [10–13]. In the UK, much work has used the UK Biobank cohort [14–18], an ongoing prospective cohort involving over 500,000 people since 2006 [19, 20]. These studies indicate that PM_2.5_, PM_10_, NO_2_, NO*_x_* may be risk factors for dementia, including AD and VAD. However, there remain unanswered questions that need addressing to allow effective translation of findings into public health policies. First, it is unclear the extent of spatial confounding of reported associations due to correlations between air pollution and area-level covariates, including socioeconomic position (SEP) and noise pollution. Second, given the extended etiological time frame for developing dementia (years to decades) [10, 11], reported associations may be sensitive to the length of the exposure time window considered, but few studies have considered longer windows (e.g., ≥ 5 years) [17, 21]. Third, findings are inconsistent with respect to which air pollutant is more relevant, partly due to their high spatial and temporal correlation. Finally, the specific effect of NO has been largely ignored, as studies have usually used NO*_x_* instead, combining NO_2_ and NO.

In this study, we aimed to rigorously investigate both individual and compound effects of PM_10_, PM_2.5_, PM*_coarse_*, PM*_abs_*, NO_2_, and NO on the risk of all-cause dementia, AD, and VAD. To do so, we triangulated several complementary approaches. We controlled for area-level covariates to minimise bias originating from spatially correlated confounders at a local level. We used variability of detected associations across UK Biobank recruitment centres to assess plausible causality: because neurotoxic effects of air pollution should be fairly constant across locations, high variability would likely indicate the presence of residual bias due to spatially correlated confounders at regional level. We then investigated which pollutants may be more harmful by using Lasso regression with multi-pollutant models, a technique suited for variable selection with highly correlated predictors. With regards to temporal considerations, we addressed the potential bias due to the length of exposure to air pollution, by comparing results using 1 or 5 years as minimum exposure time. We further evaluated the risk of bias in our analyses by triangulation with positive and negative control outcome analyses. Lastly we investigated possible interactions between air pollution exposure and SEP on risk of dementia, to identify potential vulnerable subgroups of people.

## 2 Methods

### 2.1 Study Design and Population

The UK Biobank is an ongoing prospective cohort study involving over 500,000 participants aged 40-69 years recruited between 2006 and 2010 from one of the 22 recruitment centres across England, Wales, and Scotland at baseline [19, 20]. Participants’ sociodemographic, lifestyle, clinical, genetic, and biochemical information has been collected through questionnaires, physical measurements, sampling assays, genotyping, and linked health records at baseline and throughout follow-up time. For more details on this cohort, please see [19, 20].

### 2.2 Dementia Cases

Dementia cases were algorithmically defined based on the combination of information from baseline self-reports and linked data from hospital admissions and death registries [22]. Thus, dates of dementia incidence span a time period that goes from before baseline up to now. The diagnosis of dementia incidence from linked data was based on the International Classification of Diseases codes (ICD 9/10). The date of diagnosis was equal to the earliest recorded date of dementia occurrence, irrespective of the source. Participant’s outcome could be defined as either AD (n=2679) or VAD (n=1338); all-cause dementia (n=6141) included both, in addition to other dementia types (e.g., unspecified dementia). Because we considered air pollution exposure up to baseline, we excluded participants whose dementia was detected before the baseline assessment (2006-2010) (Figure 1).

**Fig. 1.**
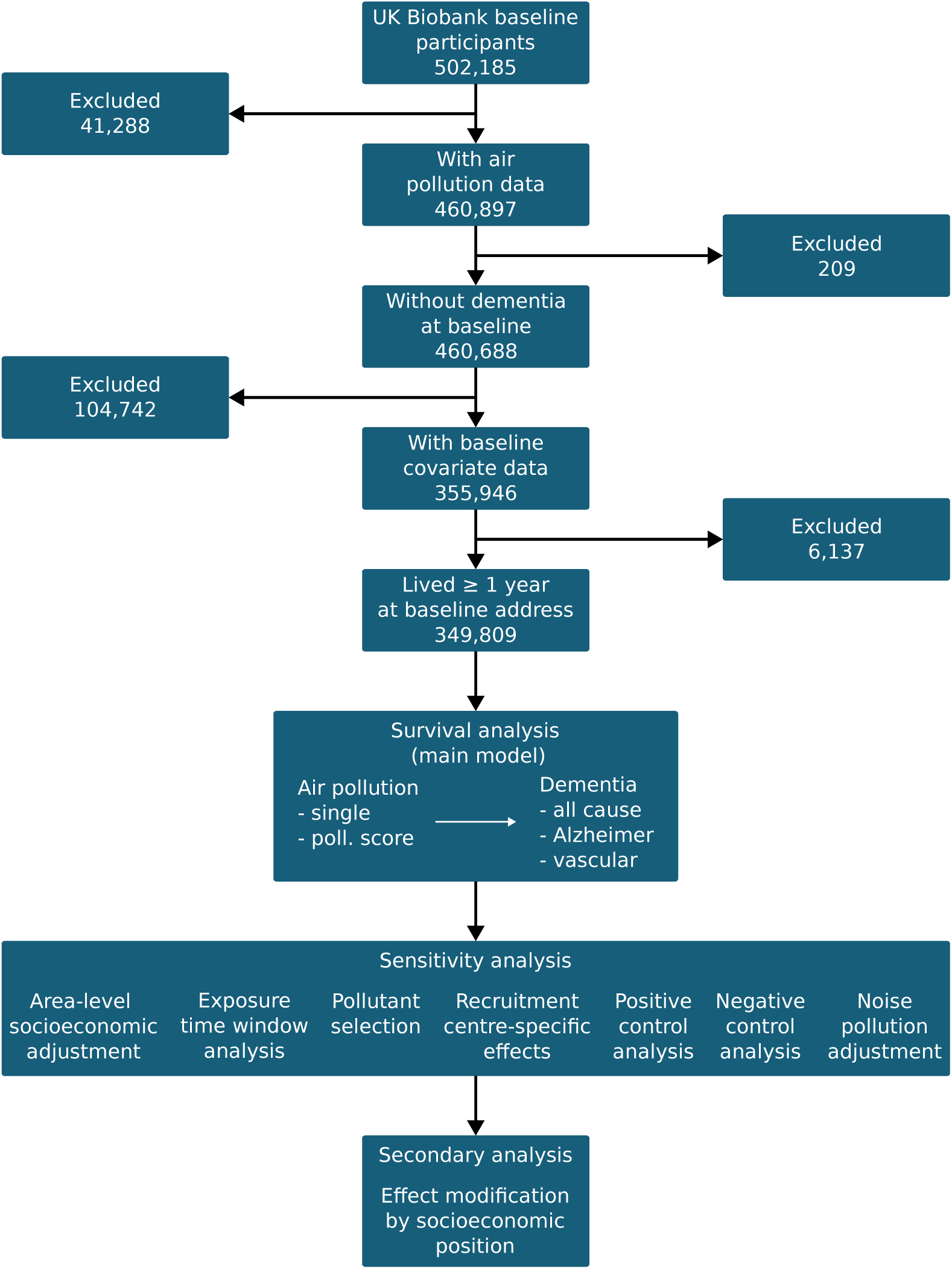
Analysis flow chart

### 2.3 Air Pollution Exposure

UK Biobank provides estimates of air pollution exposure at the residential address of participants in 2010, for an area covering up to 400 km from Greater London. We assumed relatively stable pollution concentrations over years and considered 2010 data a valid proxy for previous years’ exposures. Air pollutants for which information is present include PM_10_, PM_2.5_, PM*_coarse_*, PM*_abs_*, NO_2_ and NO (see Supplementary Information for more detail). Here we encoded air pollution exposure as both continuous variables scaled by interquartile range (IQR), and categorical variables as quartiles (see Table 1 in Supplementary Information).

**Table 1:** Distribution of air pollution exposure quartiles. Percentage of participants assigned to the 1*^st^*, 2*^nd^*, 3*^rd^* and 4*^th^* air pollution exposure quartiles in UK Biobank and within groups with and without all-cause dementia, AD, and VAD. The percentages were computed after applying exclusion criteria shown in Figure 1.

| Exposure | Overall | all-cause dementia |  | Alzheimer's |  | Vascular dementia |  |
| --- | --- | --- | --- | --- | --- | --- | --- |
|  |  | no | yes | no | yes | no | yes |
|  | 349,809 | 343,668 | 6,141 | 347,130 | 2,679 | 348,471 | 1,338 |
| **pm<sub>2.5</sub>** |  |  |  |  |  |  |  |
| 1st | 25.4% | 25.5% | 23.4% | 25.4% | 23.4% | 25.4% | 23.7% |
| 2nd | 25.2% | 25.2% | 25.5% | 25.2% | 25.3% | 25.2% | 27.0% |
| 3rd | 25.3% | 25.3% | 26.3% | 25.3% | 25.9% | 25.3% | 26.0% |
| 4th | 24.1% | 24.1% | 24.9% | 24.1% | 25.5% | 24.1% | 23.3% |
| **pm<sub>coarse</sub>** |  |  |  |  |  |  |  |
| 1st | 25.5% | 25.5% | 25.6% | 25.5% | 26.7% | 25.5% | 24.9% |
| 2nd | 25.4% | 25.4% | 26.4% | 25.4% | 25.9% | 25.4% | 28.0% |
| 3rd | 24.9% | 24.9% | 24.8% | 24.9% | 24.1% | 24.9% | 23.2% |
| 4th | 24.2% | 24.2% | 23.3% | 24.2% | 23.4% | 24.2% | 23.9% |
| **pm<sub>abs</sub>** |  |  |  |  |  |  |  |
| 1st | 27.0% | 27.0% | 25.5% | 27.0% | 25.5% | 27.0% | 24.6% |
| 2nd | 24.4% | 24.4% | 26.0% | 24.4% | 26.2% | 24.4% | 26.5% |
| 3rd | 24.0% | 24.0% | 25.5% | 24.0% | 25.2% | 24.0% | 25.1% |
| 4th | 24.6% | 24.6% | 23.0% | 24.6% | 23.1% | 24.6% | 23.8% |
| **pm<sub>10</sub>** |  |  |  |  |  |  |  |
| 1st | 25.4% | 25.4% | 24.4% | 25.4% | 24.9% | 25.4% | 24.1% |
| 2nd | 25.3% | 25.2% | 26.7% | 25.3% | 25.7% | 25.2% | 28.6% |
| 3rd | 25.0% | 25.0% | 25.7% | 25.0% | 26.4% | 25.0% | 23.3% |
| 4th | 24.4% | 24.4% | 23.2% | 24.4% | 23.1% | 24.4% | 23.9% |
| **NO<sub>2</sub>** |  |  |  |  |  |  |  |
| 1st | 25.8% | 25.8% | 23.4% | 25.8% | 24.2% | 25.8% | 23.0% |
| 2nd | 25.1% | 25.1% | 26.9% | 25.1% | 26.6% | 25.1% | 27.4% |
| 3rd | 24.5% | 24.5% | 25.5% | 24.5% | 24.4% | 24.5% | 25.8% |
| 4th | 24.6% | 24.6% | 24.2% | 24.6% | 24.7% | 24.6% | 23.8% |
| **NO** |  |  |  |  |  |  |  |
| 1st | 25.7% | 25.8% | 23.7% | 25.8% | 23.4% | 25.8% | 21.2% |
| 2nd | 25.3% | 25.3% | 25.2% | 25.3% | 25.2% | 25.3% | 27.4% |
| 3rd | 24.6% | 24.6% | 25.7% | 24.6% | 25.5% | 24.6% | 26.0% |
| 4th | 24.4% | 24.4% | 25.3% | 24.4% | 25.9% | 24.4% | 25.5% |

In this study, we considered the individual and compound neurotoxic effects of PM_10_, PM_2.5_, PM*_coarse_*, PM*_abs_*, NO_2_, and NO. Recent studies have estimated the effect of the joint pollutants’ exposure using a score defined as the subject-specific average of log hazard ratios (HRs) obtained from single-pollutant models [18, 23], which depends on the set of covariates adjusted for in the single-pollutant analysis. To avoid this dependence, we opted to use principal component analysis (PCA)[16]: specifically, we used the first principal component, accounting for 40% of overall variability in IQR-scaled air pollutant data, as a (unitless) score for the joint exposure to PM_10_, PM_2.5_, PM*_coarse_*, PM*_abs_*, NO_2_, and NO across individuals, and called this the ‘global’ air pollution score. Thus, a participant with a low global score could still be greatly exposed to a single pollutant, but have a low general level of exposure across pollutants. As the results from single-pollutant models suggested a negligible effect of PM_10_ and PM*_coarse_* on risk of dementia, we also performed PCA excluding these two pollutants, thus focusing on the most relevant variability in the data; this ‘restricted’ score was then preferred over the global one in all sensitivity analyses, and unless stated otherwise, pollution score refers to the ‘restricted’ score.

### 2.4 Confounding variables

Potential confounders included age, sex, ethnicity, household income, educational attainment, and population density (urban or rural area); these were included in all analyses. We additionally included Index of Multiple Deprivation (IMD), noise pollution and recruitment centres in sensitivity analyses. See Supplementary Information for definition of these variables and Direct Acyclic Graphs (DAG, Figure 1 in Supplementary Information). As the IMD is not comparable across countries (England, Scotland and Wales), we restricted all analyses to participants with the English IMD (∼ 95% of all participants) (Figure 1). All results are from complete case analyses, that is, participants with missing data in any variables were excluded; see Supplementary Information for more details.

### 2.5 Statistical Analysis

We assumed that putative effects of air pollution exposure during the baseline period are constant over the future risk of dementia; we therefore conducted Cox regression analyses to investigate the association between air pollution and risk of all-cause dementia, AD, and VAD. In the primary analysis, we adjusted for age, sex, ethnicity, household income, educational attainment and population density. For the exposure, in separate models, we used single air pollutants (coded either continuously, with median as reference, or categorically, with 1*^st^* quartile as reference) and air pollution score (both ’global’ score and ’restricted’ score). In all sensitivity analyses, we only used the ‘restricted’ air pollution score.

In all Cox regression analyses, we used the time from baseline as a time unit, with follow-up period ending with dementia incidence, loss to follow-up, death or end of study, whichever came first. The end of study dates used in these analyses were 31*^st^* of October 2022 for England, 31*^st^* of August 2022 for Scotland and 31*^st^* of May 2022 for Wales ^1^. We tested the proportionality hazard assumption using Schoenfeld residuals plots, and we found that age, sex, income, PM_10_, PM_2.5_, PM*_abs_* and NO_2_ appeared to violate the proportionality assumption (*p <* 0.05); however, this was likely due to the large sample size, and the distribution of residuals appeared to be flat over time (Figure 14-17 in Supplementary Information).

We conducted the following sensitivity and secondary analyses:

#### 2.5.1 Multiple deprivation index

Due to correlations in spatial distribution of air pollution and socioeconomic position, it is possible that a backdoor (i.e., noncausal) path between pollution and dementia exists through socioeconomic position. To block this pathway, we additionally adjusted for area-level socioeconomic position, by adjusting for IMD; IMD was included in all subsequent analyses. We used IMD as opposed to the Townsend deprivation index (TDI), which is more widely used in UK Biobank studies. Whereas TDI is a score of material deprivation based on four household variables (unemployment, car and home ownership, household overcrowding) [24], IMD integrates area-level deprivation, at a Lower layer Super Output Area level, in seven distinct domains, including income, employment, health/disability, education/training, barriers to housing/services, environment and crime: thus, IMD can return a much more detailed picture of deprivation^2^.

#### 2.5.2 Long term exposure

As the effects of air pollution on the brain are likely cumulative, it is important to take into account the exposure (timeframe) window. We examined whether associations between air pollution and dementia are sensitive to the length of exposure to air pollution by using 1 (1-year cutoff) versus 5 (5-year cutoff) years as a minimal time window for pollution exposure, as indicated by the time lived at baseline residence; we chose 5 years because it gives a good tradeoff between minimal length of (reported) pollution exposure and number of participants additionally dropped from the analyses (∼ 15%). These analyses were adjusted for IMD.

#### 2.5.3 Multi-pollutant analysis

Because it is possible that only a few pollutants may be the driving risk factors for dementia, we applied Lasso (L1) penalisation to the estimation of coefficients in the multi-pollutant model. This is a method for variable selection that aims to address the problem of collinearity, which here arises due to spatial correlations between pollutants (see Supplementary Table 21). The L1 penalty was selectively applied to the estimation of air pollutants coefficients, excluding covariates (see Supplementary Information). For this analysis, we used both 1- and 5-years cutoff and adjusted for IMD.

#### 2.5.4 Recruitment centre analysis

Putative causal effects of air pollution on dementia are expected to be fairly constant across locations; alternatively, variability across locations may be due to unaccounted bias. We thus assessed the spatial variability of the air pollution effect on dementia, by including random effects for UK Biobank recruitment centres in the Cox regression, with random effects encompassing a recruitment centre-specific intercept and air pollution slope; in this context, the variability in slope is of particular interest. This analysis was possible because participants appeared to be highly clustered around recruitment centres (Figures 18, 19 in Supplementary Information), which can therefore be used as proxy for geographical location. For this analysis we used both 1- and 5-years cutoff and adjusted for IMD.

#### 2.5.5 Positive control analysis

We performed positive control analysis with time to COPD incidence as an outcome. This analysis aims at detecting bias in the data, by checking if an existing association could be detected in the population of interest. We chose COPD because there is strong evidence that air pollution is a risk factor for this condition [25], and the set of confounders is likely to be similar as for dementia (see [26]; excluding lifestyle factors as mentioned in our discussion). As for the dementia analysis, we performed Cox regression using the incidence of algorithmically defined COPD cases as an outcome, and excluded participants whose COPD incidence occurred before baseline assessment. For this analysis we used both 1- and 5-years cutoff and adjusted for IMD.

#### 2.5.6 Negative control analysis

We performed negative control analysis with frequency of oily fish intake as a negative control outcome. This analysis aims to detect the presence of bias in our analysis after controlling for confounders, as any observed association between exposure and outcome would be likely due to selection bias or residual confounding. Fish intake frequency was chosen because it is unlikely to be directly caused by air pollution exposure, while the set of confounders are plausibly the same to those involved for dementia. Oily fish intake frequency was assessed through baseline self-report questionnaires (see Supplementary Information), and was encoded ordinally. We assessed the association between exposure to air pollution and frequency of oily fish intake using ordinal logistic regression, with and without adjustment for IMD.

#### 2.5.7 Noise pollution

We additionally controlled for noise pollution exposure. Noise and air pollution are likely to be influenced by some common factors, for example vicinity to roads or topography of a city; on the other hand, noise pollution could be a risk factor for dementia incidence [27], and could thus confound the association between air pollution and dementia. For this analysis we used both 1- and 5-years cutoff and adjusted for IMD.

#### 2.5.8 Effect modification by IMD

From a public health policy standpoint, it is relevant to know whether neurotoxic effects of air pollution change with socioeconomic position. We therefore estimated in a secondary analysis the interactions between air pollution exposure and IMD on the additive and multiplicative scale: as a measure for interactions on the additive scale we used the relative excess risk due to interaction (RERI), whereas for interactions on the multiplicative scale we used the product term in the linear model of the Cox regression [28].

## 3 Results

### 3.1 Participant characteristics

After exclusion criteria were applied, the total number of participants available for the main analysis was 349,809 (Figure 1). For these participants, Table 1 shows the overall distribution of air pollution exposure quartiles, as well as the distributions for participants grouped by presence or absence of the different dementia outcomes. Similarly, Table 2 shows the characteristics of the same participants in terms of confounders used for the analyses.

**Table 2:** Characteristics of the confounders. Categorical variables are described in terms of percentage, continuous variables are described with their mean and standard deviation, the latter in parenthesis. These characteristics are shown for the UK Biobank population and within groups with and without all-cause dementia, AD, and VAD. The characteristics were computed after applying restriction criteria shown in Figure 1. The unit for education is years of education, for noise pollution is decibel. Please see Supplementary Information for more information on covariate coding.

| Characteristics | Overall | all-cause dementia |  | Alzheimer's |  | Vascular dementia |  |
| --- | --- | --- | --- | --- | --- | --- | --- |
|  |  | no | yes | no | yes | no | yes |
|  | 349,809 | 343,668 | 6,141 | 347,130 | 2,679 | 348,471 | 1,338 |
| **age** | 56.3 (8.1) | 56.2 (8.0) | 64.1 (4.8) | 56.3 (8.1) | 64.5 (4.3) | 56.3 (8.1) | 64.7 (4.2) |
| **sex** |  |  |  |  |  |  |  |
| female | 52.6% | 52.7% | 43.9% | 52.6% | 48.4% | 52.6% | 36.7% |
| **ethnicity** |  |  |  |  |  |  |  |
| white | 95.0% | 94.9% | 96.5% | 95.0% | 96.9% | 95.0% | 96.7% |
| other | 5.0% | 5.1% | 3.5% | 5.0% | 3.1% | 5.0% | 3.3% |
| **education** | 14.2 (4.9) | 14.2 (4.9) | 12.4 (5.0) | 14.2 (4.9) | 12.2 (5.0) | 14.2 (4.9) | 12.0 (4.8) |
| **income** |  |  |  |  |  |  |  |
| inc. <18k | 22.7% | 22.3% | 44.7% | 22.6% | 44.7% | 22.6% | 48.1% |
| inc. 18-31k | 25.6% | 25.6% | 30.7% | 25.6% | 32.0% | 25.6% | 30.3% |
| inc. 31-52 | 26.1% | 26.2% | 16.0% | 26.1% | 15.5% | 26.1% | 14.9% |
| inc. 52-100k | 20.2% | 20.4% | 7.1% | 20.3% | 6.3% | 20.3% | 5.7% |
| inc. >100k | 5.4% | 5.4% | 1.5% | 5.4% | 1.5% | 5.4% | 1.0% |
| **population density** |  |  |  |  |  |  |  |
| urban | 85.6% | 85.6% | 87.1% | 85.6% | 86.6% | 85.6% | 88.2% |
| rural | 14.4% | 14.4% | 12.9% | 14.4% | 13.4% | 14.4% | 11.8% |
| **deprivation (q)** |  |  |  |  |  |  |  |
| 1st | 25.7% | 25.7% | 23.4% | 25.7% | 23.0% | 25.7% | 23.5% |
| 2nd | 25.4% | 25.4% | 24.1% | 25.4% | 25.3% | 25.4% | 21.7% |
| 3rd | 24.8% | 24.9% | 23.5% | 24.8% | 24.5% | 24.8% | 23.3% |
| 4th | 24.1% | 24.0% | 29.0% | 24.1% | 27.3% | 24.1% | 31.5% |
| **noise pollution** | 56.0 (4.3) | 56.0 (4.3) | 55.9 (4.2) | 56.0 (4.3) | 55.9 (4.2) | 56.0 (4.3) | 56.0 (4.4) |
| **lost follow-up** |  |  |  |  |  |  |  |
| no | 99.8% | 99.8% | 99.9% | 99.8% | 100.0% | 99.8% | 99.9% |
| **lost death** |  |  |  |  |  |  |  |
| no | 91.8% | 92.5% | 51.4% | 92.1% | 53.2% | 92.0% | 42.6% |

### 3.2 Air pollution exposure and dementia association

For continuously encoded exposures, we found some evidence of a negative effect of PM_2.5_ (HR=1.06, 1.03-1.10), PM*_abs_* (HR=1.04, 1.01-1.08), NO_2_ (HR=1.08, 1.04-1.12) and NO (HR=1.03, 1.01-1.06), but not of PM*_coarse_* (HR=0.99, 0.97-1.02) and PM_10_ (HR=1.01, 0.99-1.04), on all-cause dementia risk (Figure 2). We found similar results, but with stronger evidence, when considering AD as an outcome: PM_2.5_ (HR=1.10, 1.04-1.15), PM*_abs_* (HR=1.06, 1.02-1.11), NO_2_ (HR=1.11, 1.05-1.17) and NO (HR=1.05, 1.01-1.09) (Supplementary Figure 2). In contrast, there was little evidence of an effect of any air pollutants on VAD, as HR were closer to 1 and confidence intervals were wider (Supplementary Figure 3). Overall, PM_2.5_ and NO_2_ were the pollutants with stronger evidence of a neurotoxic effect: an increase of 1 unit in the IQR scale (1.27 *µg/m*^3^ and 9.86 *µg/m*^3^) corresponded to a 6% and 8% increase of risk in all-cause dementia, and a 10% and 11% increase of risk in AD, respectively. For NO_2_, this is equivalent to a 16% and 22% higher risk of all-cause dementia and AD, respectively, for a person living in large and densely populated city like London compared to small coastal city like Plymouth in Devon (∼ 19*µg/m*^3^ difference in NO_2_ concentration^3^). Evidence was weaker for PM*_abs_* and NO, and little for PM*_coarse_* and PM_10_.

**Fig. 2.**
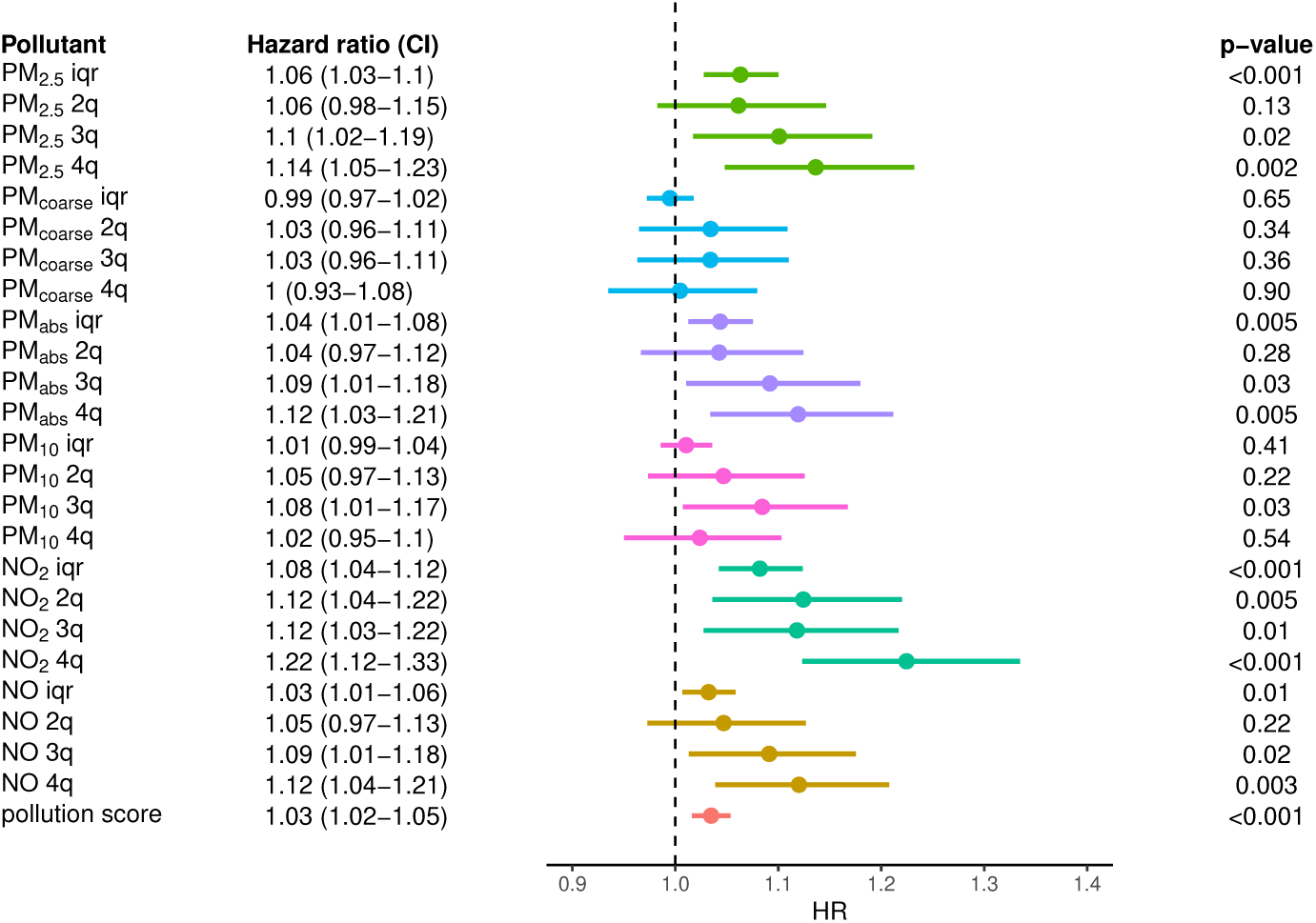
Effect of air pollution on all-cause dementia. From left to right, we show the pollution exposure with the associated HR and confidence interval, the forest plot and p-value of the effect estimate (Wald test); ‘iqr’ and ‘*n^th^*q’ refer to the continuous (scale by IQR) and discrete exposures (*n^th^* quartile), respectively. The models were adjusted for age, sex, ethnicity, educational attainment, income and population density.

When using air pollution exposures encoded as quartiles, results were consistent with the above. In general, the effect sizes across pollutants for all-cause dementia and AD showed a dose-response trend, with the 4*^th^* quartile showing the highest concentration of air pollution has the strongest impact on dementia.

As for the air pollution score, we found evidence of an adverse effect on all-cause dementia (HR=1.03, 1.02-1.05, Figure 2), AD (HR=1.05, 1.02-1.08, Supplementary Figure 2) but not VAD (HR=1.02, 0.98-1.07, Supplementary Figure 3). We found similar results when using the global pollution score as the exposure (not shown); however, for both all-cause dementia (HR=1.006, 1.002-1.010) and AD outcomes (HR=1.009, 1.003-1.014) evidence was weaker, likely due to the inclusion of PM*_coarse_* and PM_10_ score. In conclusion, higher joint exposure to (IQR-scaled) PM_2.5_, PM*_abs_*, NO_2_, and NO appeared to have a risk-inducing effect on all-cause dementia and AD risk, although evidence was weaker than that for individual PM_2.5_ and NO_2_ effects.

### 3.3 Sensitivity Analyses

#### 3.3.1 Inclusion of multiple deprivation index

Adjusting for multiple deprivation index (IMD) resulted in less evidence of a neurotoxic effect for all pollutants (Figure 3). Specifically, for continuous pollutant exposures and all-cause dementia, we found little evidence of an association for all air pollutants, except NO_2_ (HR=1.04, 1.00-1.08). With AD as an outcome (Supplementary Figure 4), evidence was also reduced (although it was still stronger than for all-cause dementia), such that there was some evidence of an association for PM_2.5_ (HR=1.07, 1.01-1.13), NO_2_ (HR=1.07, 1.01-1.14) and the pollution score (HR=1.04, 1.01-1.07). With VAD as an outcome (Supplementary Figure 5), we found little evidence of an association for all air pollutants. A similar pattern was observed using air pollution quartiles as exposure.

**Fig. 3.**
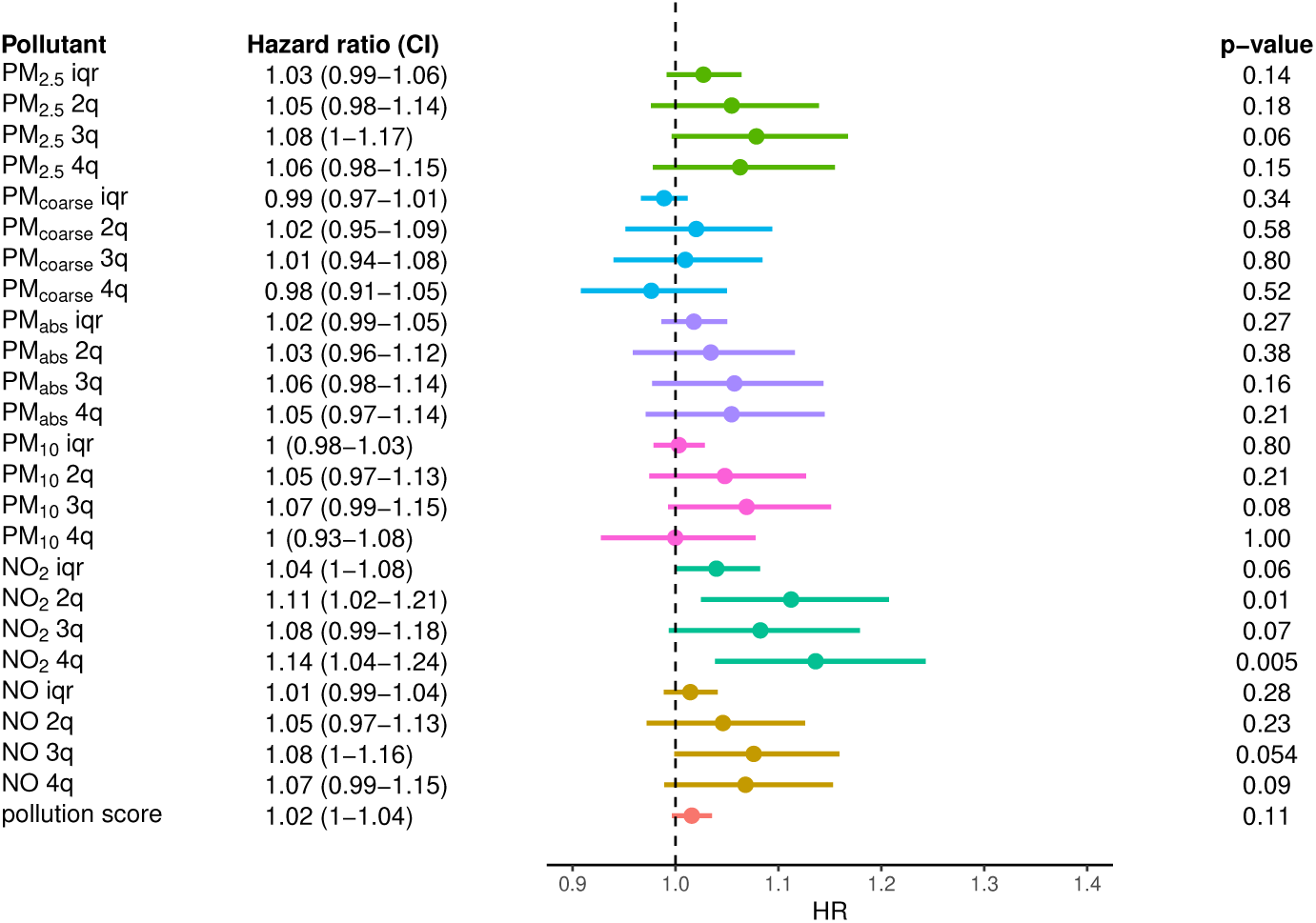
Effect of air pollution on all-cause dementia. From left to right, we show the pollution exposure with the associated HR and confidence interval, the forest plot and p-value of the effect estimate (Wald test); ‘iqr’ and ‘*n^th^*q’ refer to the continuous (scale by IQR) and discrete exposures (*n^th^* quartile), respectively. The models were adjusted for age, sex, ethnicity, educational attainment, income, population density and deprivation index.

Altogether, IMD seemed to strongly confound the association between air pollution and dementia, such that after adjustment for it only NO_2_ appeared consistent as a risk factor: an increase by 9.86 *µg/m*^3^ in concentration corresponded to a 4% and 7% increase in risk of all-cause dementia and AD, respectively.

#### 3.3.2 Long term exposure

We then repeated the analysis adjusted for IMD, but selectively including participants who lived ≥ 5 years (5-year cutoff) instead of ≥ 1 year (1-year cutoff) at baseline residence. We found stronger evidence of a neurotoxic effect for most air pollutants. With all-cause dementia as an outcome (Figure 4), there was some evidence of a positive association for continuously encoded PM_2.5_ (HR=1.04, 1.00-1.08), NO_2_ (HR=1.06, 1.02-1.11) and pollution score (HR=1.02, 1.00-1.04). As before, evidence was stronger with AD as an outcome (Supplementary Figure 6): PM_2.5_ (HR=1.08, 1.02-1.14), NO_2_ (HR=1.09, 1.02-1.16) and pollution score (HR=1.04, 1.01-1.07). Again, we found little evidence of an association between any air pollutant and VAD incidence (Supplementary Figure 7). We found analogous results when using air pollution exposure quartiles (Figure 4, Supplementary Figures 6 and 7).

**Fig. 4.**
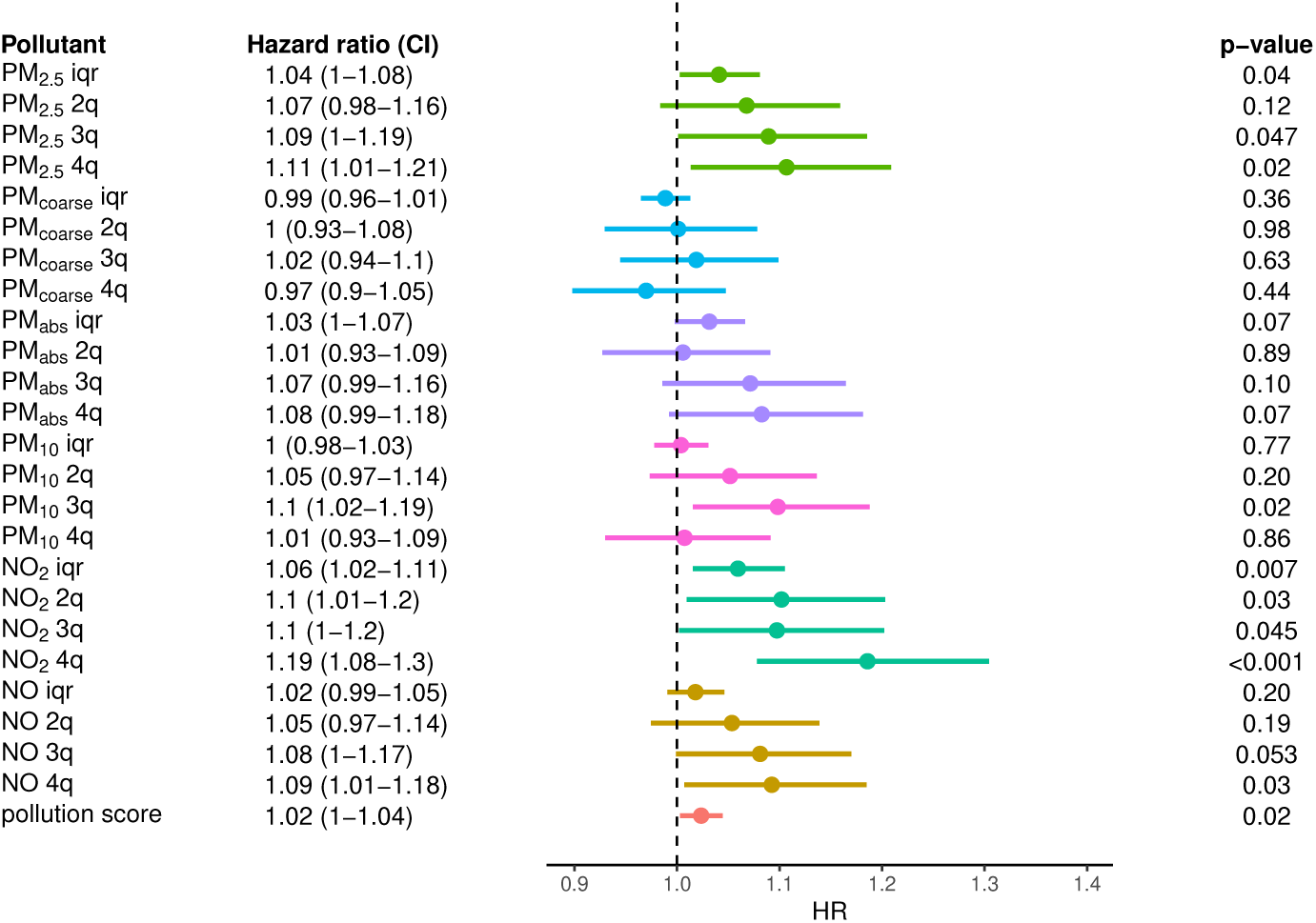
Effect of air pollution on all-cause dementia. From left to right, we show the pollution exposure with the associated HR and confidence interval, the forest plot and p-value of the effect estimate (Wald test); ‘iqr’ and ‘*n^th^*q’ refer to the continuous (scale by IQR) and discrete exposures (*n^th^* quartile), respectively. The models were adjusted for age, sex, ethnicity, educational attainment, income, population density and deprivation index. Additionally, here we excluded participants who lived less that 5 years (instead of 1 year) at baseline address.

Thus, after accounting for IMD as well as longer exposure time, 1 IQR increase in PM_2.5_ and NO_2_ was associated with a 4% and 6% increase in all-cause dementia risk (compared to 3% and 4% with 1-year cutoff), and 8% and 9% increase in AD (compared to 7% and 7% with 1-year cutoff).

#### 3.3.3 Variable selection analysis

The results above point to NO_2_ and possibly PM_2.5_ as main risk factors for dementia among air pollutants. To better investigate which air pollutants are stronger risk factors for all-cause dementia, we applied Lasso regression to a multi-pollutant model. The results of this analysis are shown in Table 2 of the Supplementary Information. When using 5-years cutoff only NO_2_ (HR=1.0021) appeared to have an effect. Hence, this analysis suggested NO_2_ as the main risk factor for dementia.

#### 3.3.4 Recruitment centre analysis

We assessed the consistency of the association of air pollution exposure on all-cause dementia by including pollution exposure as a random effect varying across recruitment centres. The results of this analysis are shown in Figure 8 and Table 3 of the Supplementary Information. The largest average centre-specific HR was reported for NO_2_ using 5-years cutoff (mean=1.019): although in agreement with the above results, there was also considerable variability across recruitment centres (sd=0.076), suggesting little consistency of the association across England. Notably, only the HR’s for PM*_coarse_* and PM_10_ were highly concentrated around 1, suggesting no effect on dementia risk. Therefore, this analysis highlighted some difference in the recruitment centre-specific HR’s for most pollutants, potentially indicating the presence of bias at a regional level.

#### 3.3.5 Positive control

We used COPD as a positive control outcome. Accordingly, we found evidence of an association for all pollutants and COPD, with effect sizes in general much larger compared to those obtained with dementia as an outcome (Figure 9 of the Supplementary Information). Similarly to the dementia results above, adjusting for IMD (Figure 10 of the Supplementary Information) reduced the evidence of an association for all air pollutants; however, in contrast to the dementia analysis, the increase in evidence when using a baseline period ≥ 5 years was much smaller, if present (Figure 12 of the Supplementary Information). Overall, the positive control analysis lent support to our previous results, but also highlighted a possible difference in terms of etiological time window between dementia and COPD.

#### 3.3.6 Negative control

We used oily fish intake frequency as a negative control outcome. For this analysis, we focused on the comparison between models adjusted for or not adjusted for IMD. Without adjustment, we found little evidence of an effect of air pollution, although with a perceptible general tendency to increase fish frequency intake (Supplementary 4). Adjusting for IMD slightly increased the association between air pollution and higher oily fish intake for all pollutants, suggesting that including IMD in the analysis may introduce some bias.

#### 3.3.7 Control for noise pollution

We adjusted for noise pollution in addition to baseline covariates and IMD, using 5-year cutoff. Inclusion of noise pollution did not affect the results (13 in the Supplementary Information). In particular, we found some evidence of an effect of NO_2_ (HR=1.08, 1.03-1.13) on all-cause dementia: this effect is comparable to that previously obtained without inclusion of noise pollution (Figure 4), suggesting no confounding by the latter.

#### 3.3.8 Effect modification by deprivation index

We examined the modification of the effect of air pollution on dementia by IMD. We report both the effect modification on the additive and multiplicative scale in Tables 5-18 of the Supplementary Information. For all pollutants and both 1- and 5-year cutoff, there was little evidence of effect modification on both scales.

## 4 Discussion

We examined the individual and compound effect of PM_2.5_, PM_10_, PM*_coarse_*, PM*_abs_*, NO_2_ and NO on the risk of developing all-cause dementia, AD, and VAD. We addressed several factors hindering both the causal interpretation and translation in public health policies of detected effects using the UK Biobank cohort. We found NO_2_ to be the strongest risk factor for dementia, especially when focusing on AD as an outcome. Sensitivity analyses highlighted the importance of considering longer exposure periods and accounting for spatial confounding. Together, our results highlight that targeting air pollution, in particular NO_2_ levels, could inform preventive public health policies for dementia.

We found stronger evidence for an effect of PM_2.5_ and NO_2_ on all-cause dementia risk, whereas evidence was lacking for PM*_coarse_* and PM_10_. Similar results were found when treating pollution exposures as IQR-scaled continuous or categorical variables; in the latter case, being in the 4*^th^* quartile of the pollution exposure distribution represented generally the greatest risk. Lack of evidence for larger particles (PM*_coarse_* and PM_10_) is congruent with some [14, 15, 29, 30] but not all [17, 18, 31, 32] UK Biobank studies. Mixed results may be explained by differences in the analyses used, including the set of covariates adjusted for: in the present work, we excluded variables (e.g., lifestyle) whose effects on air pollution are uncertain (e.g., smoking, drinking, physical activity), as they may introduce selection bias.

Our analyses using air pollution score suggested that combined exposure to PM_2.5_, PM*_abs_*, NO_2_ and NO does not increase the risk of dementia more than individual pollutant exposures. Although it is important to examine the health effect of mixtures rather than isolated pollutants [33], these analyses, in the context of dementia incidence, speak against possible super-additive interactions between different air pollutants, at least at a coarse, population level. Instead, it is simpler to explain our results by interpreting the compound effect of pollution as a sum of single pollutants effect. This leads to the question about which air pollutant may be a stronger risk factor for dementia. Variable selection indicated NO_2_ as the driving factor, suggesting that detected associations for PM*_abs_* and NO and possibly PM_2.5_, may reflect to some extent NO_2_ concentrations. This would in turn explain the smaller effect size of the pollution score compared to that of NO_2_: the score discarded some variability associated with NO_2_ exposure in order to accommodate that from PM_2.5_, PM*_abs_*, and NO.

Compared to all-cause dementia, there was more evidence of a neurotoxic effect for all pollutants, except PM_10_ and PM*_coarse_*, with AD as an outcome. Notably, this was observed despite the fact that AD cases (n=2,679) were fewer than those for all-cause dementia (n=6,141). On the other hand, there was little evidence of an association with VAD for any pollutants. These results are in line with other studies that found stronger evidence for an effect on AD than VAD [14, 16, 34], but are also at odds with known mechanistic pathways linking air pollution to cardiovascular disease [35], and cardiovascular disease to dementia [36]. Together, this could suggest that air pollution unevenly promotes different neurodegenerative mechanisms underlying dementia subgroups, for example, by affecting protein structure and functions more than the neurovascular system via oxidative stress and inflammation. However, we also note that VAD had relatively few cases (n= 1,338) compared to AD, which could bias the analyses.

When restricting analyses to participants who had ≥ 5 years of baseline air pollution exposure (as opposed to ≥ 1 year), there was more evidence of a neurotoxic effect for pollutants except PM_10_ and PM*_coarse_*, which is consistent with findings from few other studies testing this [17, 21]. Interestingly, we did not observe the same change when using COPD as a positive control: this suggests that the specific consequences of air pollution on the nervous system may only become apparent on longer time scales, reflecting the extended etiological time window for dementia. Previous studies, both in the UK Biobank [14, 16, 18, 29, 30, 32], and outside [10, 11] have included participants with shorter reported periods of exposure, and this could have biased the results. It may be difficult to identify the optimal temporal restriction criteria to use, as this should account for the trade off with reduced sample size (∼ 15% in our sample) and possible selection bias induced. The availability of datasets with increased sample size and information about past address history are necessary to tackle these questions.

Regarding spatial confounding, we tried to assess and account for it at both the local and regional level. When adjusting for IMD, there was less evidence of an association with both all-cause dementia and AD for all air pollutants previously found harmful, to the extent that only increase in NO_2_ exposure levels appeared consistently harmful. These results 1) hint at the importance of accounting for (local) area-level SEP to avoid overconfidence in the results, and 2) highlight the robustness of evidence of the role played by NO_2_ as a risk factor for dementia. When interpreting these results, however, it is important to note that negative control analyses suggested that including IMD may also introduce some amount of bias in the results. Finally, we did not find much evidence in support of increased vulnerability to pollution based on SEP; although these results are plausible, effect modifications may still be present for subgroups of people at particular extremes of the SEP scale, which may not be well represented in the UK Biobank cohort [37].

At a regional level, we assessed the spatial variability of detected associations across UK Biobank recruitment centres. Using random effect models, we detected negligible variability in the recruitment centre-specific HRs for PM*_coarse_* and PM_10_, which were tightly clustered around 1, thus providing further evidence of their negligible association with dementia. On the other hand, for all other pollutants and pollution score, variability was an order of magnitude higher, with standard deviations on the HR scale between 3-7%, and several centre-specific HR’s below 1. Whilst we again found the greatest average effect for NO_2_, this heterogeneity hints at the presence of unmeasured, systematic differences in participants attending various recruitment centres, for example due to regional stratification of socioeconomic and healthcare related variables, or due to different selection bias mechanisms operating across England.

There are several limitations that may undermine our conclusions and whose impact could not be tested here. First, outdoor measures of air pollution levels used in this study do not comprehensively capture individual exposure, neither in space nor in time. Indeed, exposure computed at residential address does not account for indoor levels as well as outdoor levels experienced during commuting or at work. Similarly, concentrations for 2010 can only return noisy estimates of long-term exposure because of changes in pollutant concentrations over time, for example, due to the introduction of the clean air zones. Previous work has found the temporal approximation valid when comparing results using time-varying exposure levels [16]; nonetheless, more precise measures may improve estimates of the cumulative effects of air pollution on dementia and enable investigation of relevant exposure time windows, as mentioned above. Second, measurement of time of dementia onset can be biased, as it depends on factors such as frequency of use of the healthcare system and presence of comorbidities, which in turn may cause spurious associations. Third, the classification of dementia, even in its major subtypes, is highly problematic and debated; thus, results indicating a differential effect of air pollution on dementia subtypes need to consider these issues. Finally, our results may have limited generalisability; this is a common problem of studies on air pollution and dementia, which mostly include people from high-income countries [38–44]. Although mechanisms underlying the effect of air pollution are shared, the context and other population-specific differences may also play a critical role.

Despite the above limitations, this work clearly strengthens evidence supporting the role of air pollution, in particular NO_2_, as a key risk factor for dementia, and advocates for actions aimed at improving air quality management. The estimated NO_2_ and PM_2.5_ annual air pollution exposure levels for participants included in this study well exceeded annual air quality guideline levels set by the World Health Organisation (NO_2_ 10 *µg/m*^3^, PM_2.5_ 5*µg/m*^3^) [45], with even minimum levels surpassing these thresholds (NO_2_ 12.93*µg/m*^3^, PM_2.5_ 8.170*µg/m*^3^) (see Table 20 in Supplementary Material); this highlights the importance of decreasing emissions, concentrations and exposure to air pollution [46]. Because the main anthropogenic source of NO_2_ is fossil fuel combustion for heating, power generation, and vehicle exhaust [47], addressing emissions requires policies promoting the use of cleaner energy sources. At the same time, as most participants lived in or around urban centres, which are the areas with highest pollution concentration (see Figures 18 in Supplementary Information); these results also emphasise the importance of a type of urban planning that, within a general framework of increased quality of life and healthcare, aims at reduced the concentration of and exposure to air pollution, especially NO_2_.

## 5 Conclusion

The present work adds evidence in support of the role of air pollution in increasing the risk for dementia, while addressing some key questions and issues in the field. We reported neurotoxic effects for various air pollutants, and in particular PM_2.5_ and NO_2_, even after comprehensively adjusting for socioeconomic deprivation. Notably, there was strongest evidence for an involvement of NO_2_, which we propose as a key target for public health policies. Additionally, we found that air pollution was a stronger risk factor for AD than VAD; this differential effect, in turn, warrants more research on the mechanisms underlying neurotoxic effects of air pollution. Given the smaller sample size of VAD (compared to AD and all-cause dementia) studies with larger samples of VAD are also needed. Finally, our results also highlight the strong necessity to consider appropriately extended time windows for air pollution exposure, at least within the range of 5 years, in studies of dementia.

## Data Availability

Data can be made available upon request to the UK Biobank data access team.

https://www.ukbiobank.ac.uk/

## Declarations

### 5.1 Supplementary information

The article has an accompanying supplementary file.

## 5.2 Acknowledgements

This research has been conducted using the UK Biobank Resource under application number 80288, and uses data provided by patients and collected by the NHS as part of their care and support.

## 5.5 Ethics approval and consent to participate

All participants consented to link their electronic health records (EHR) from the National Health Service (NHS) to the study database. The UK Biobank study has received ethical approval by the North West Multicentre Research Ethics Committee.

## 5.6 Consent for publication

The UK Biobank Science team consented to the publication of this article.

## 5.7 Data availability

Data can be made available upon request to the UK Biobank data access team.

## 5.8 Material availability

Not applicable

## 5.9 Clinical trial number

Not applicable

## 5.10 Code availability

Code to reproduce the analyses is available at https://github.com/ensorpalacios/apoll-dem-analysis.

## 5.11 Author contribution

E.R.P, G.J.G and K.T. contributed to the conception, analysis and writing of the problem; C.Y.S, L.J.W., L.T.N, E.L.A., C.S. and P.G.K. contributed to the analysis and writing; D.B and D.M. contributed to the writing.

## Footnotes

1 please see https://biobank.ndph.ox.ac.uk/ukb/exinfo.cgi?src=Data_providers_and_dates

2 please see https://biobank.ndph.ox.ac.uk/ukb/refer.cgi?id=6810 for more information

3 confront UUNN_LOND_999 and UUNN_PLYM_001 in https://uk-air.defra.gov.uk/data/exceedance?f_exceedence_id=S3&f_year_start=2023&f_year_end=2024&f_group_id=31&f_region_reference_id=5&f_parameter_id=NO2-BA&f_sub_region_id=9999&f_output=screen&action=exceedance3&go=Submit

