## Supplementary Information for "Dissecting the effect of long-term exposure to air pollution on risk of dementia in UK Biobank"

**2 Missing data** **4**

**3 Alzheimer's disease and vascular dementia outcomes** **5**

**4 Variable selection analysis** **11**

**5 Recruitment centre random effects** **12**

**6 Positive control analysis** **14**

**7 Negative control analysis** **18**

**8 Control for noise pollution** **19**

**9 Effect modification by deprivation level** **20**

**10 Proportionality assumption check** **24**

**11 Air pollutants characteristics** **28**

### 1 UK Biobank variables

Number within parentheses indicates the UK Biobank data field for the associated variables.

#### 1.1 Outcomes

We used algorithmically defined outcomes for all cause dementia (UK Biobank field 42018), Alzheimer's disease (AD, 42020), vascular dementia (VAD, 42022), and chronic obstructive pulmonary disorder (COPD; 42016) data; ICD 9/10 codes can be found at `https://biobank.ndph.ox.ac.uk/showcase/refer.cgi?id=460`. Oily fish intake frequency data were collected at baseline with the question 'How often do you eat oily fish?' (1329), to which participants could respond with 'Never', 'Less than once a week', 'Once a week', '2-4 times a week', '5-6 times a week', 'Once or more daily', ' Do not know' and 'Prefer not to answer'; we encoded '2-4 times a week', '5-6 times a week' and 'Once or more daily' as '$>$ 2 per week', because the latter two responses accounted for only $\sim$ 1% or the responses, whereas 'Do not know' and 'Prefer not to answer' were considered as missing. In order to preserve the direction of the association detected between air pollution, index of multiple deprivation (IMD) and dementia incidence, when instead using fish intake frequency, the order of the encoding was as follows: '$>$ 2 per week' $<$ 'once a week' $<$ '$<$ once a week' $<$ 'never'.

#### 1.2 Exposure

We used estimates of individual residential address exposure to $PM_{2.5}$ (24006), $PM_{absorbance}$ (24007), $PM_{2.5-10}$ (24008), $PM_{10}$ (24005), $NO_2$ (24003) and $NO_x$ (24004) for the year 2010. Exposure estimates were obtained by linking each participant's residential address to the annual average concentrations of air pollution derived at a $100m^2$ resolution from land use regression (LUR) models, as part of the European Study of Cohorts for Air Pollution Effects (ESCAPE) [1, 2]. In that study, $NO_x$ was measured using a passive samplers with a filter for both NO and $NO_2$; thus, NO measures could be obtained by subtracting $NO_2$ from $NO_x$ levels [3]. In the UK, the output of the LUR models were tested using data from air pollution monitors placed in the London/Oxford and Manchester areas, hence estimates are valid up to 400 km from Greater London. Consequently, all participant with addresses outside this area have missing data, and were therefore excluded from the analysis (Figure 1 in the manuscript). Air pollution data from the ESCAPE project have been shown to match well with data derived from UK air information resources [4, 5].

| | q.0% | q.25% | q.50% | q.75% | q.100% | IQR |
|---|---|---|---|---|---|---|
| pm25 | 8.17 | 9.28 | 9.92 | 10.55 | 21.25 | 1.27 |
| pmcoarse | 5.57 | 5.84 | 6.10 | 6.62 | 12.82 | 0.78 |
| pmabs | 0.83 | 0.99 | 1.13 | 1.30 | 4.57 | 0.31 |
| pm10 | 11.78 | 15.23 | 16.02 | 16.98 | 30.65 | 1.75 |
| no2 | 12.93 | 21.31 | 26.07 | 31.17 | 108.49 | 9.86 |
| no | -0.28 | 11.61 | 15.94 | 20.53 | 160.06 | 8.92 |

Table 1: Quartiles and inter-quartile range (IQR) of air pollutants. Units of measures are in $\mu g/m^3$.

#### 1.3 Covariates

Information about age (21022), sex (31), ethnicity (21000), educational attainment (6138), household income (738) and recruitment centre (54) was collected through baseline questionnaires or based on location of assessment centre. Age was encoded as a continuous variable, sex and recruitment centre as categorical. For ethnicity, we categorised participants as 'white' if they replied 'White', 'British', 'Irish', 'Any other white background', or as 'other' for everything else. For educational attainment, we created an educational score based on years of education following Okbay[6], except that we capped the score for national vocational qualification (NVQ), higher national diploma (HND) and higher national certificate (HNC) at 19, so that is was less than 20 for college. Additionally, participants who replied 'do not know', 'prefer not to say', or did not reply were assigned the average score, whereas participants who replied 'none of the above' were assigned the minimum score of 7. For household income, we categorised participants as '$< 18,000$', '18,000-30,999', '31,000-51,999', '52,000-100,000', and '$> 100,000$'.

Other covariates were computed based on the participants home postcode, including population density (20118), IMD for England (26410) and average 24-hour sound level in dB of noise pollution (24024). Information about

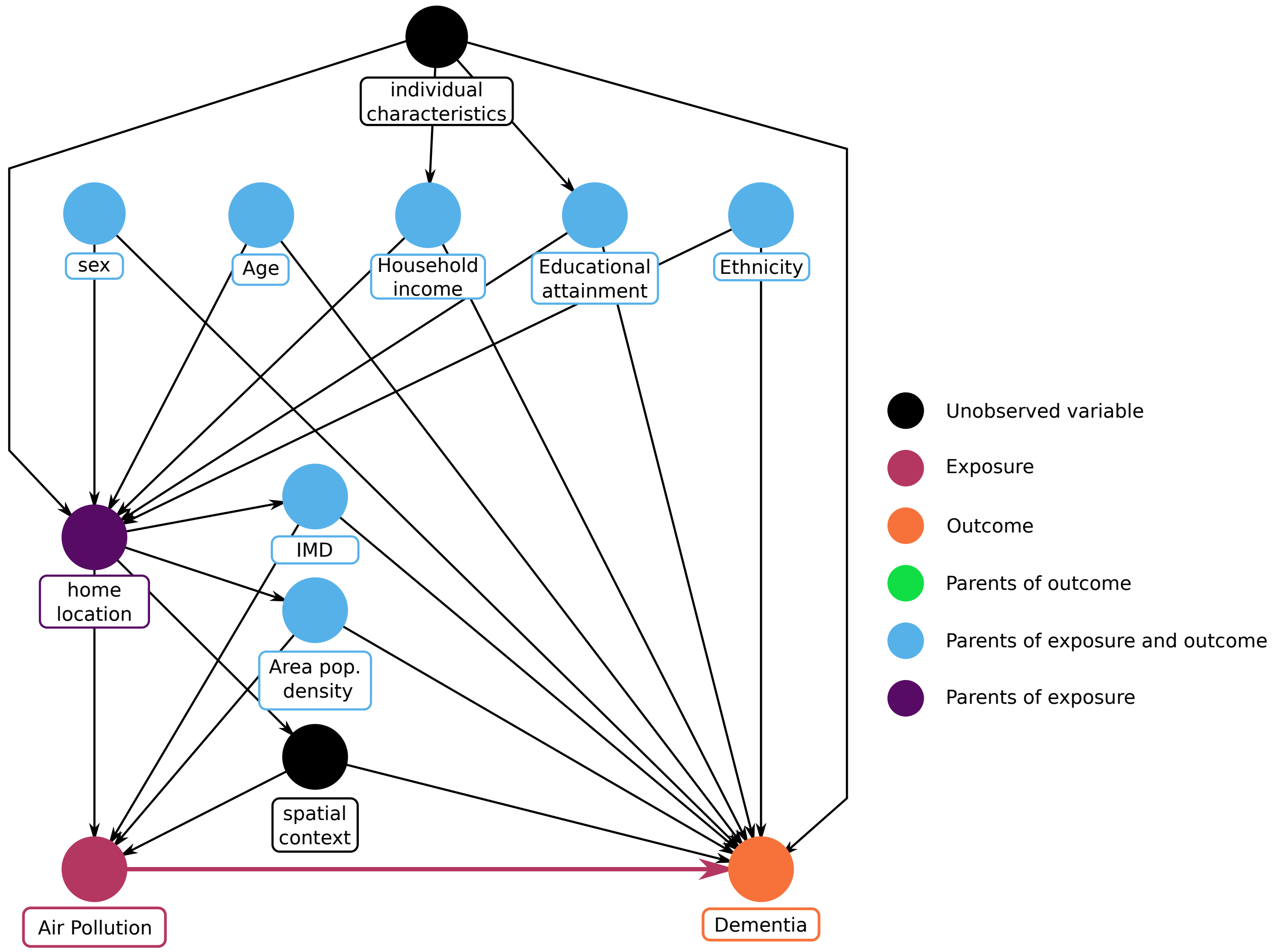

Figure 1: Direct acyclic graph. Index of multiple deprivation (IMD)

population density was derived from data generated from the 2001 census from the Office of National Statistics, and participants were categorised as 'urban' or 'rural', reflecting the meaning assigned to each code in the UK Biobank showcase (`https://biobank.ndph.ox.ac.uk/showcase/coding.cgi?id=91`). The IMD is a compound measure of deprivation at a small area level reflecting deprivation in seven distinct domains, including income, employment, health/disability, education/training, barriers to housing/services, environment and crime. Here we encoded deprivation categorically as quartiles, with the $1^{st}$ quartile as reference. Noise estimates for year 2009 were modelled using a Common Noise Assessment Methods in Europe (CNOSSOS-EU) noise model [7]; noise pollution was encoded as a continuous variable.

### 2 Missing data

In the UK Biobank study, missing data for air pollution (n=41286) exposure are only due to the participants living outside the area within which LUR model estimates are considered valid (up to 400 km from Great London); this mechanisms is likely to cause missing completely at random data in the context of assessing the effect of air pollution exposure on dementia incidence. On the other hand, the mechanisms underlying missingness for the other baseline covariates, and in particular for income, which accounted for most missing data (86%), possibly depend on the covariates themselves (missing not at random mechanism). Because of this, we chose to perform a complete case analysis, which in this scenario would give unbiased results, assuming no other unobserved variable is associated with missing mechanisms [8]. After exclusion due to missing pollution exposure and pre-existing dementia, missing data for ethnicity, educational attainment, income, and population density were respectively 2400, 4570, 71696 and 4230; for English IMD, after excluding participants with IMD for Scotland and Wales, the number of missing values was 12447, most of which were due to missing Government's data (`https://biobank.ndph.ox.ac.uk/showcase/refer.cgi?id=6810`). Among participants with pollution and IMD data, 95% had an English IMD, and $\sim 5\%$ had a Scottish or Welsh IMD. The number of participants dropped because of missing pollution or covariate values was 146013, leaving 356115 out of 502128 participants.

### 3 Alzheimer's disease and vascular dementia outcomes

We used Cox regression to assess whether air pollution affects time to dementia occurrence. Figure 2 and 3 show the results for the pollution score and single-pollutant models with AD and VAD as an outcome, respectively. These models were adjusted for age, sex, ethnicity, educational attainment, income, and population density. Air pollution exposures were encoded either continuously, scaled by inter-quartile range (iqr), or categorically, as quartiles (2q, 3q, 4q); for categorical variables, the reference was the $1^{st}$ quartile (not shown). For the (restricted) pollution score models, the exposure was the first principal component capturing variability in exposure to $PM_{2.5}$, $PM_{abs}$, $NO_2$, and NO.

Figure 4 and 5 show the results for AD and VAD, respectively, when using models additionally adjusted for IMD. Figure 6 and 7 show the results for AD and VAD, respectively, when additionally excluding participants who lived less than 5 years (instead of 1 year) at baseline address.

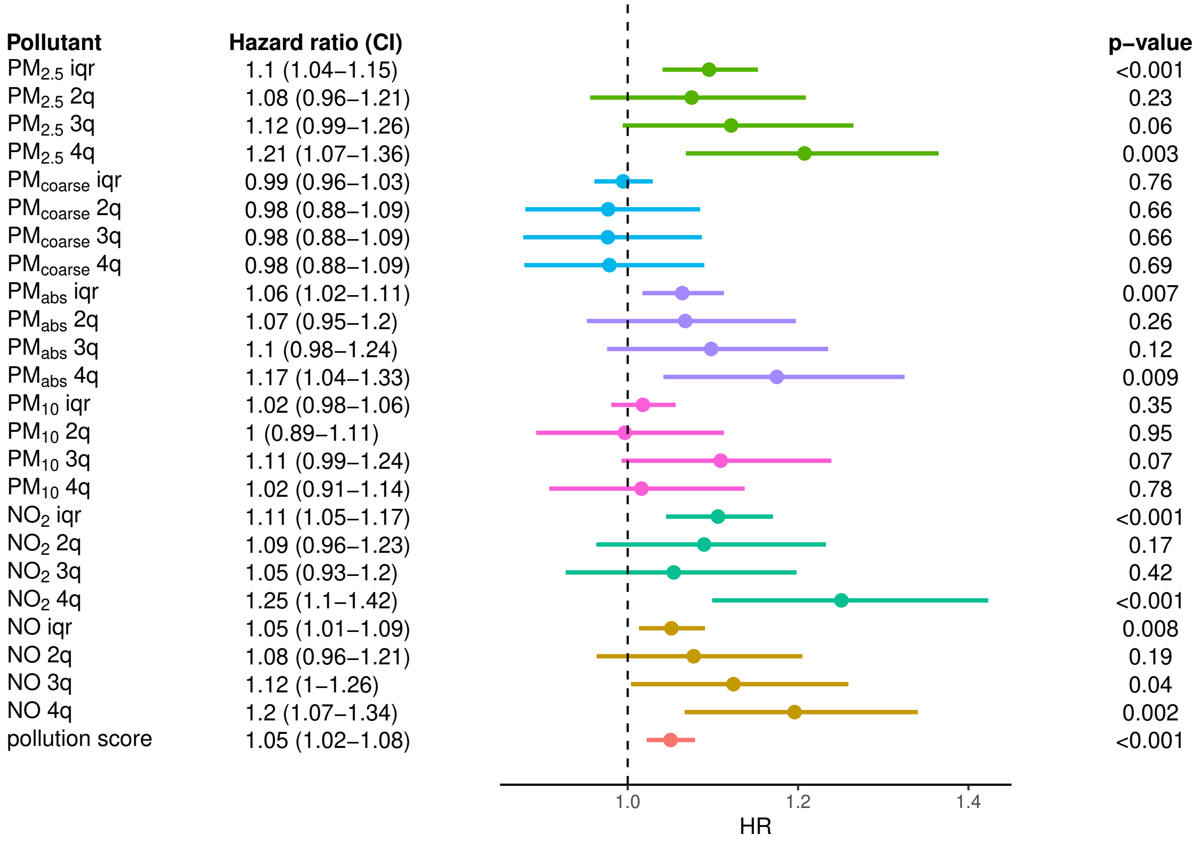

Figure 2: Effect of air pollution on AD. From left to right, for each air pollution exposure we show its associated hazard ratio and confidence interval, the forest plot and p-value of the effect estimate (Wald test); 'iqr' and '$n^{th}$q' refer to the continuous (scale by IQR) and discrete exposures ($n^{th}$ quartile), respectively. The models were adjusted for age, sex, ethnicity, educational attainment, income, and population density.

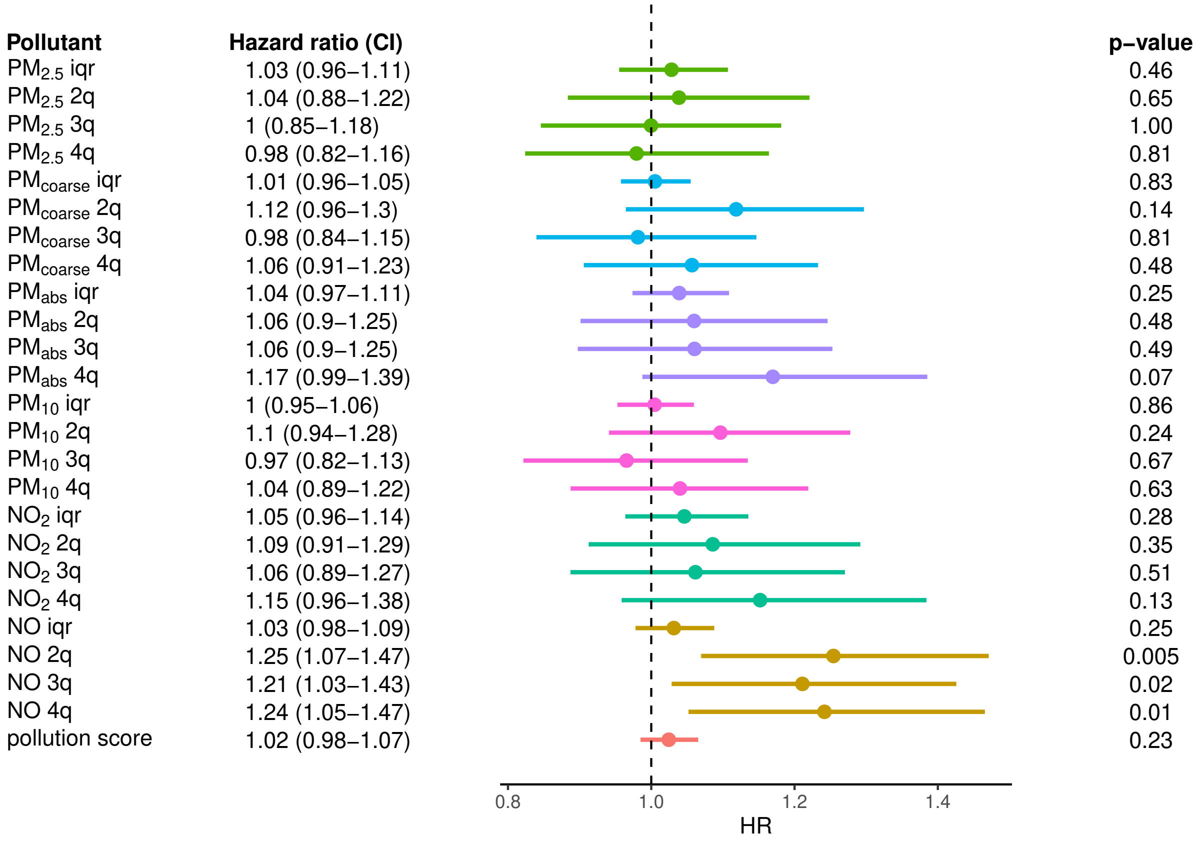

Figure 3: Effect of air pollution on VAD. From left to right, for each air pollution exposure we show its associated hazard ratio and confidence interval, the forest plot and p-value of the effect estimate (Wald test); 'iqr' and '$n^{th}$q' refer to the continuous (scale by IQR) and discrete exposures ($n^{th}$ quartile), respectively. The models were adjusted for age, sex, ethnicity, educational attainment, income, and population density.

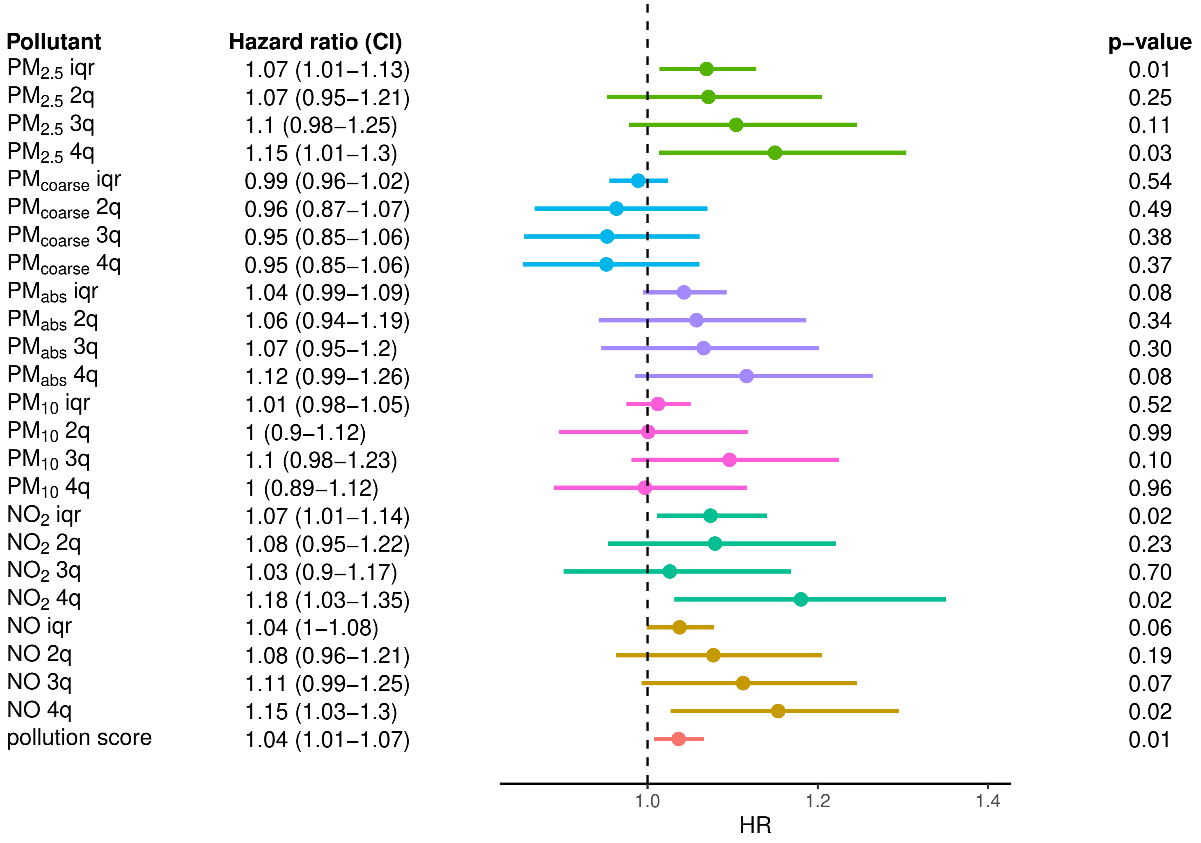

Figure 4: Effect of air pollution on AD. From left to right, we show the pollution exposure with the associated hazard ratio and confidence interval, the forest plot and p-value of the effect estimate (Wald test); 'iqr' and '$n^{th}$q' refer to the continuous (scale by IQR) and discrete exposures ($n^{th}$ quartile), respectively. The models were adjusted for age, sex, ethnicity, educational attainment, income, population density, and IMD.

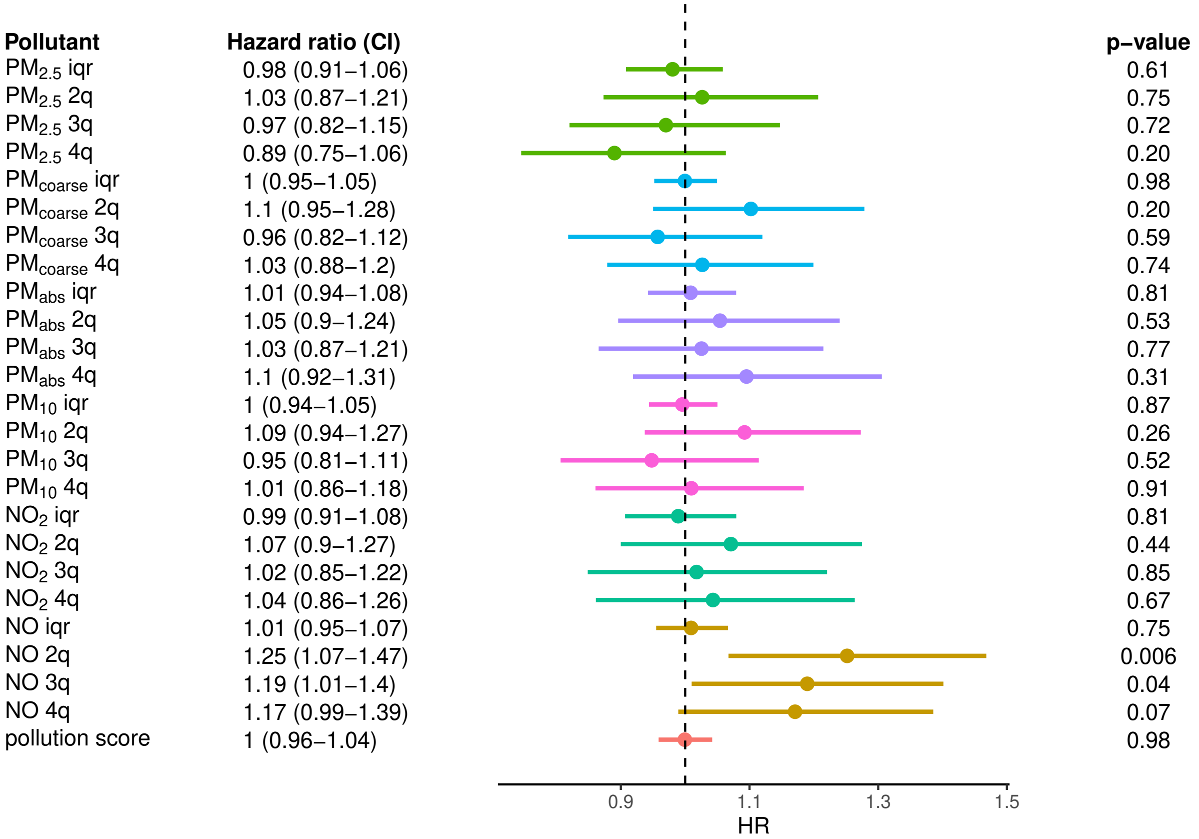

Figure 5: Effect of air pollution on VAD. From left to right, we show the pollution exposure with the associated hazard ratio and confidence interval, the forest plot and p-value of the effect estimate (Wald test); 'iqr' and '$n^{th}$q' refer to the continuous (scale by IQR) and discrete exposures ($n^{th}$ quartile), respectively. The models were adjusted for age, sex, ethnicity, educational attainment, income, population density, and IMD.

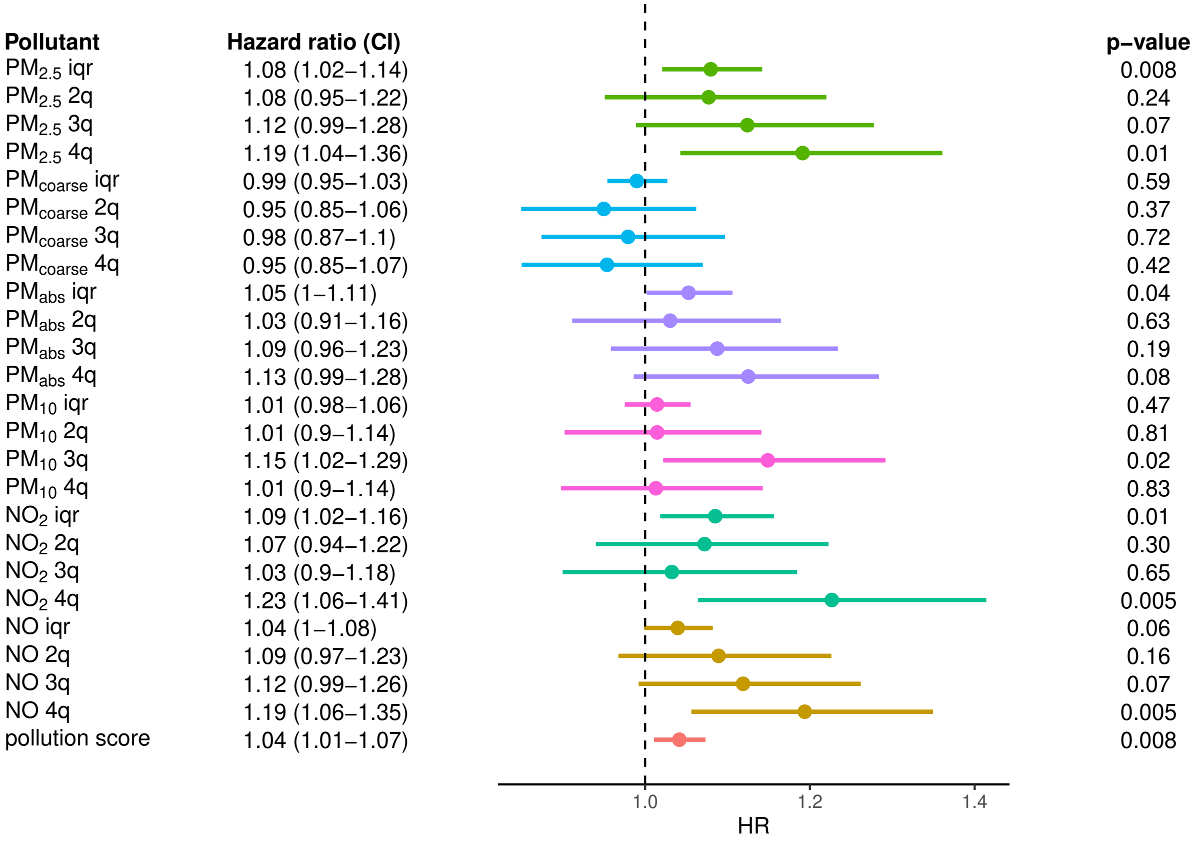

Figure 6: Effect of air pollution on AD. From left to right, we show the pollution exposure with the associated hazard ratio and confidence interval, the forest plot and p-value of the effect estimate (Wald test); 'iqr' and '$n^{th}$q' refer to the continuous (scale by IQR) and discrete exposures ($n^{th}$ quartile), respectively. The models were adjusted for age, sex, ethnicity, educational attainment, income, population density, and IMD. Additionally, here we excluded participants who lived less than 5 years (instead of 1 year) at baseline address.

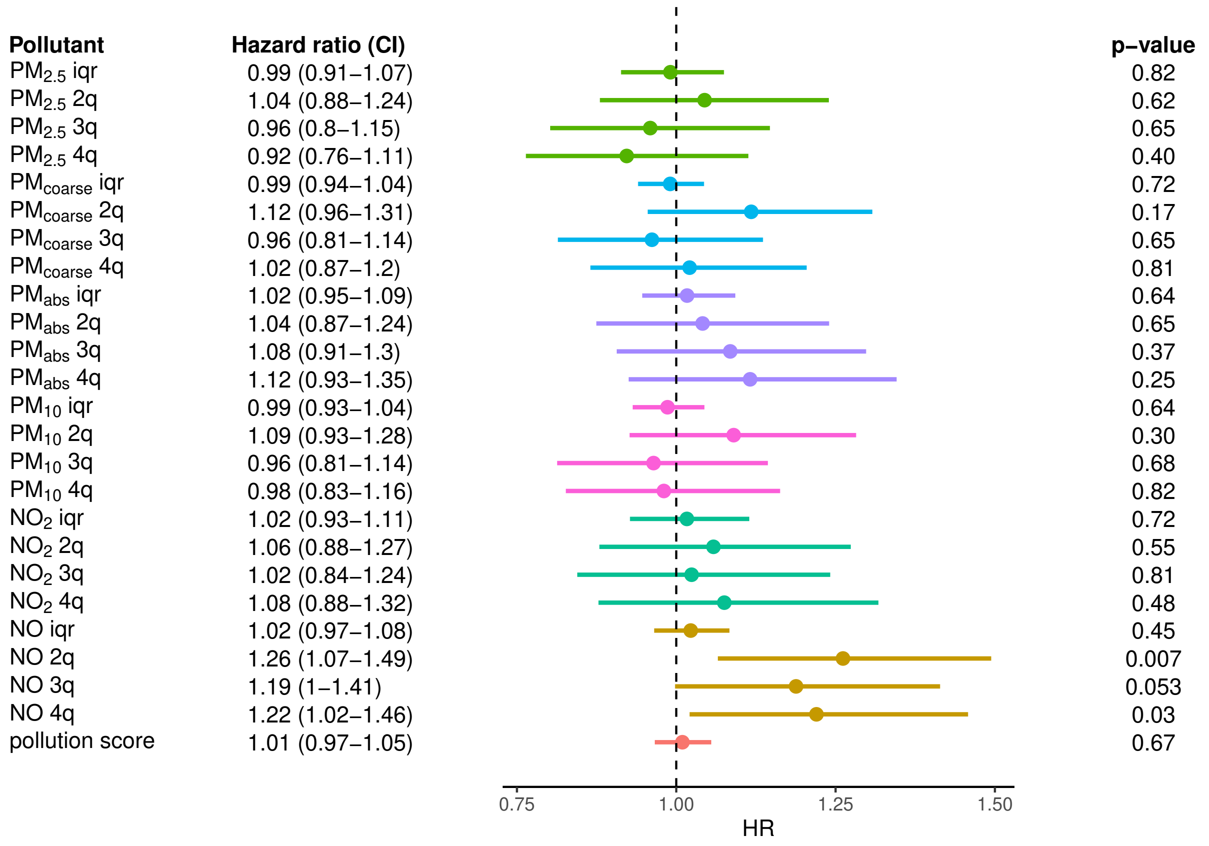

Figure 7: Effect of air pollution on VAD. From left to right, we show the pollution exposure with the associated hazard ratio and confidence interval, the forest plot and p-value of the effect estimate (Wald test); 'iqr' and '$n^{th}$q' refer to the continuous (scale by IQR) and discrete exposures ($n^{th}$ quartile), respectively. The models were adjusted for age, sex, ethnicity, educational attainment, income, population density, and IMD. Additionally, here we excluded participants who lived less than 5 years (instead of 1 year) at baseline address.

### 4 Variable selection analysis

In order to identify the most relevant air pollutants for dementia risk, we used Cox regression with a Lasso (L1) penalisation in models including all air pollutants. This penalisation constraints the coefficients towards 0, while setting a subset of them to 0, enforcing sparsity. We ran the analysis adjusting for IMD, in addition to baseline covariates, and by using both 1 and 5 years as a threshold for time at residential address. We implemented 10-fold cross-validated regularised Cox regression with L1 penalty and C-index as a loss function using the package glmnet. The L1 penalty was only applied to coefficients associated to air pollutants, thus it did not affect estimation of covariate coefficients. The air pollution coefficients from the selected model are shown in Table 2.

| | $\geq$ 1 year | $\geq$ 5 year |
|---|---|---|
| $PM_{2.5}$ | 1.0003 | 1.0000 |
| $PM_{coarse}$ | 1.0000 | 1.0000 |
| $PM_{abs}$ | 1.0000 | 1.0000 |
| $PM_{10}$ | 1.0000 | 1.0000 |
| $NO_2$ | 1.0015 | 1.0021 |
| NO | 1.0000 | 1.0000 |

Table 2: Variable selection for air pollutants. The table includes pollutant-specific coefficients for a model either excluding participants with less than 1 year or less than 5 years at baseline residence. The strength of the L1 penalisation is a free parameter, which here was chosen based on 10-fold cross-validated the C index, an evaluation metrics for survival models. The models were adjusted for age, sex, ethnicity, educational attainment, income, population density, and IMD.

### 5 Recruitment centre random effects

Random effect models included a centre-specific intercept and slope for the effect of air pollution on dementia. We ran the analysis adjusting for IMD, in addition to baseline covariates, and by using both 1 and 5 years as a threshold for time at residential address; for each model, after exclusion criteria were applied, the number of recruitment centres were 18 and 17, respectively. Figure 8 and table 3 display the distribution and standard deviations of the slopes, respectively.

| | sd | sd (HR scale) | HR mean |
|---|---|---|---|
| $PM_{2.5} \geq 1$ year | 0.038 | 1.0387 | 1.005 |
| $PM_{2.5} \geq 5$ years | 0.059 | 1.0608 | 1.013 |
| $PM_{coarse} \geq 1$ year | 0.006 | 1.0060 | 1.000 |
| $PM_{coarse} \geq 5$ years | 0.008 | 1.0080 | 1.000 |
| $PM_{abs} \geq 1$ year | 0.041 | 1.0419 | 1.002 |
| $PM_{abs} \geq 5$ years | 0.048 | 1.0492 | 1.006 |
| $PM_{10} \geq 1$ year | 0.005 | 1.0050 | 1.000 |
| $PM_{10} \geq 5$ years | 0.003 | 1.0030 | 1.000 |
| $NO_2 \geq 1$ year | 0.063 | 1.0650 | 1.010 |
| $NO_2 \geq 5$ years | 0.076 | 1.0790 | 1.019 |
| NO $\geq$ 1 year | 0.041 | 1.0419 | 1.005 |
| NO $\geq$ 5 years | 0.042 | 1.0429 | 1.006 |
| pollution score $\geq$ 1 year | 0.029 | 1.0294 | 1.003 |
| pollution score $\geq$ 5 years | 0.034 | 1.0346 | 1.007 |

Table 3: Distribution of centre-specific effect measures. In the table we show standard deviation of the pollution-specific random effects (left) and the mean of the hazard ratios over recruitment centres (right). In addition, we show the hazard ratio associated with an increase of the pollution coefficient by 1 standard deviation above 0 (middle). We display the results when excluding participants who lived either less than 1 or 5 years at baseline address.

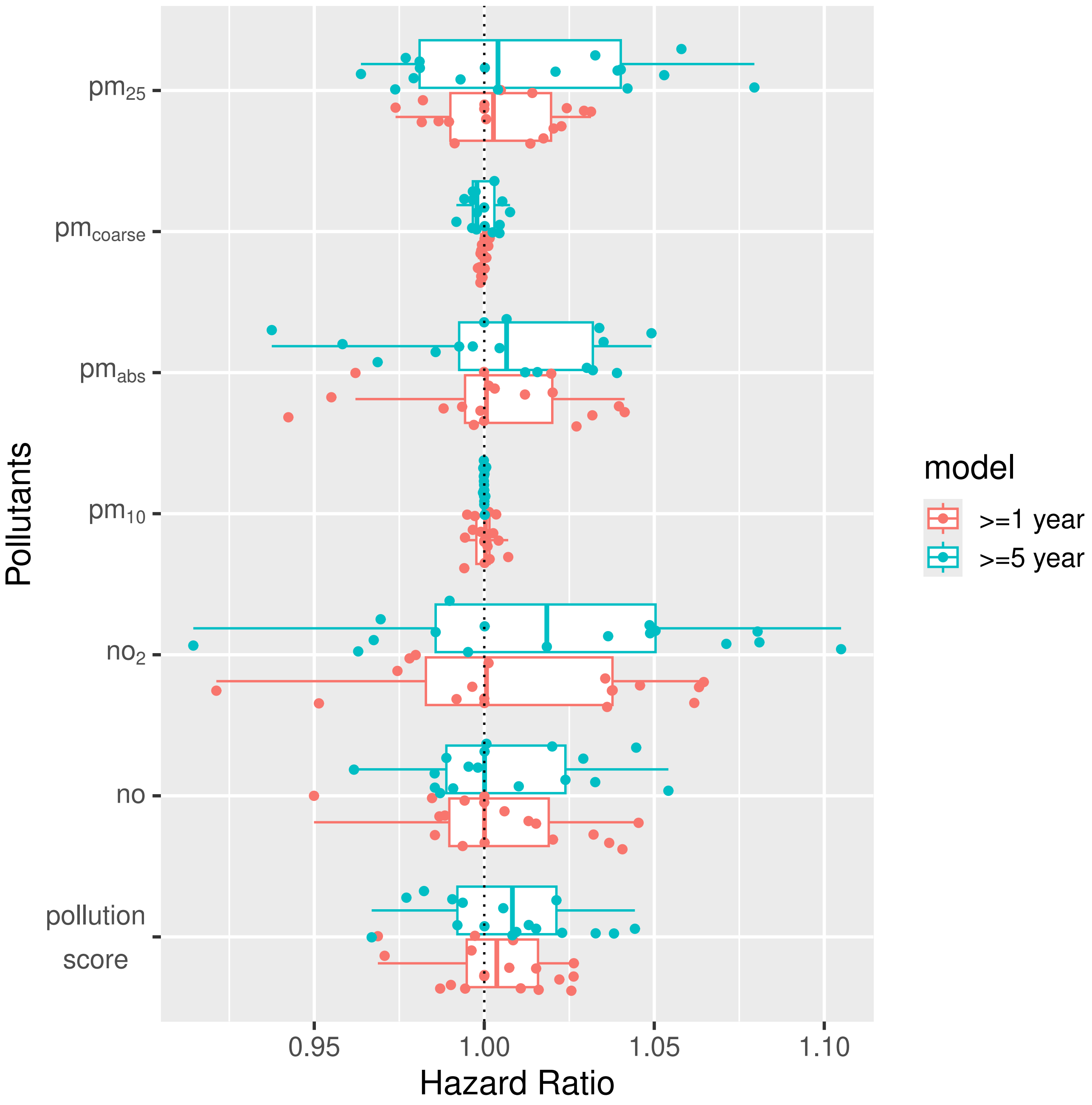

Figure 8: Pollutant-specific effects on dementia across recruitment centres. The boxplots include the $1^{st}$, $2^{nd}$ and $3^{rd}$ quartiles, plus the minimum and maximum values within 1.5*IQR; superimposed on the boxplots are the centre-specific estimates (dots). For each air pollutant we ran two analyses, excluding participants who lived either less than 1 year (red) or less than 5 years (cyan) at baseline residence; for the second analysis there was one less recruitment centre, for a total of 17 (Swansea was dropped due to lack of participants). The models were adjusted for age, sex, ethnicity, educational attainment, income, population density, and IMD.

### 6 Positive control analysis

We used Cox regression to assess whether air pollution affects time to COPD occurrence. Figure 9, 10 and 11 show the results for the pollution score and single-pollutant models respectively without IMD, with IMD as a covariate and using a 5-year cutoff for time at residential address.

Figure 12 shows the distribution of changes in HR across air pollutants when using 5 vs 1 year cutoff for time at baseline address, for all-cause dementia and COPD as an outcome.

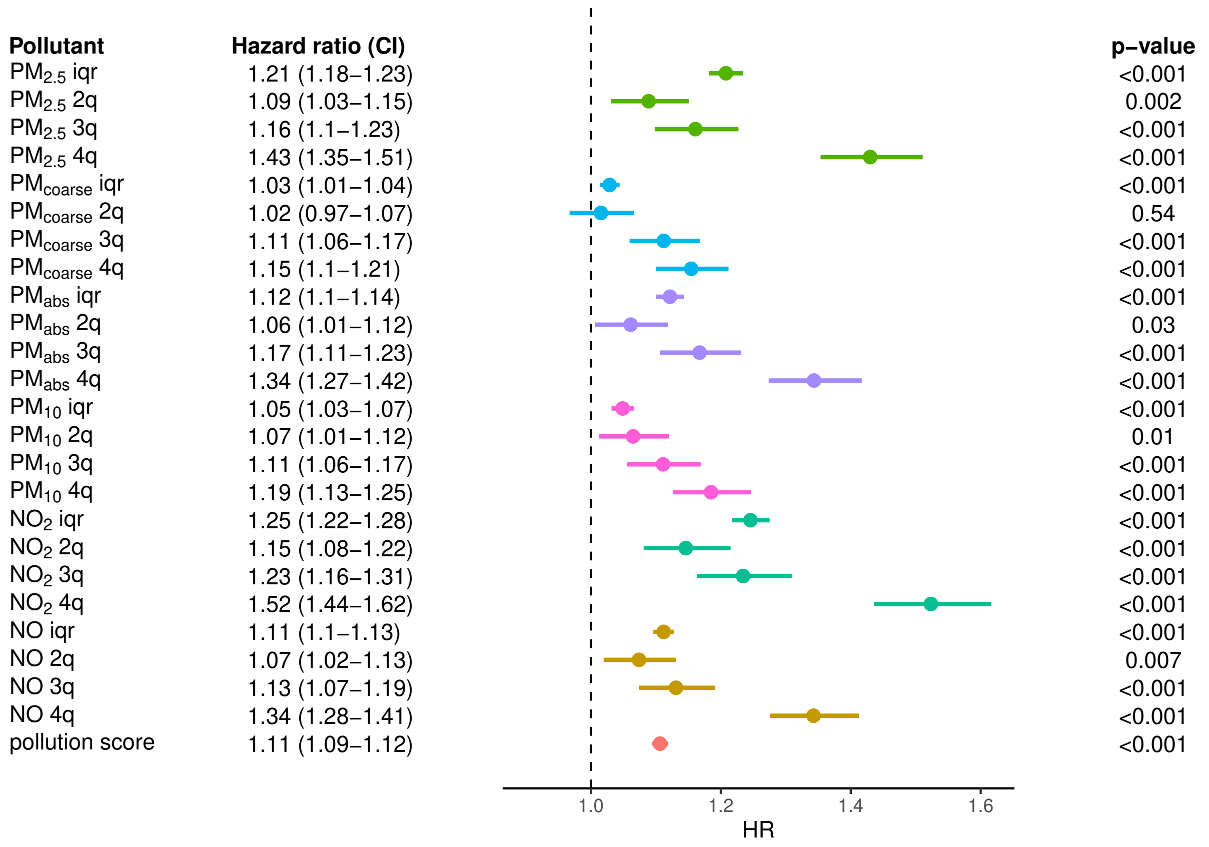

Figure 9: Effect of air pollution on COPD. From left to right, for each air pollution exposure we show its associated hazard ratio and confidence interval, the forest plot and p-value of the effect estimate (Wald test); 'iqr' and '$n^{th}$q' refer to the continuous (scale by IQR) and discrete exposures ($n^{th}$ quartile), respectively. The models were adjusted for age, sex, ethnicity, educational attainment, income, and population density.

| Pollutant | Hazard ratio (CI) | p−value |
|---|---|---|
| $PM_{2.5}$ iqr | 1.1 (1.08–1.13) | <0.001 |
| $PM_{2.5}$ 2q | 1.07 (1.01–1.13) | 0.02 |
| $PM_{2.5}$ 3q | 1.08 (1.02–1.14) | 0.006 |
| $PM_{2.5}$ 4q | 1.18 (1.12–1.25) | <0.001 |
| $PM_{coarse}$ iqr | 1.01 (1–1.03) | 0.18 |
| $PM_{coarse}$ 2q | 0.97 (0.92–1.02) | 0.19 |
| $PM_{coarse}$ 3q | 1.02 (0.97–1.07) | 0.36 |
| $PM_{coarse}$ 4q | 1.05 (1–1.1) | 0.049 |
| $PM_{abs}$ iqr | 1.04 (1.02–1.06) | <0.001 |
| $PM_{abs}$ 2q | 1.01 (0.96–1.07) | 0.63 |
| $PM_{abs}$ 3q | 1.03 (0.98–1.09) | 0.27 |
| $PM_{abs}$ 4q | 1.1 (1.04–1.16) | 0.001 |
| $PM_{10}$ iqr | 1.03 (1.01–1.04) | 0.002 |
| $PM_{10}$ 2q | 1.07 (1.02–1.13) | 0.009 |
| $PM_{10}$ 3q | 1.06 (1.01–1.11) | 0.03 |
| $PM_{10}$ 4q | 1.1 (1.05–1.16) | <0.001 |
| $NO_2$ iqr | 1.11 (1.08–1.14) | <0.001 |
| $NO_2$ 2q | 1.09 (1.03–1.16) | 0.004 |
| $NO_2$ 3q | 1.09 (1.03–1.16) | 0.005 |
| $NO_2$ 4q | 1.19 (1.12–1.27) | <0.001 |
| NO iqr | 1.06 (1.05–1.08) | <0.001 |
| NO 2q | 1.07 (1.01–1.13) | 0.01 |
| NO 3q | 1.08 (1.02–1.14) | 0.004 |
| NO 4q | 1.17 (1.11–1.23) | <0.001 |
| pollution score | 1.05 (1.04–1.07) | <0.001 |

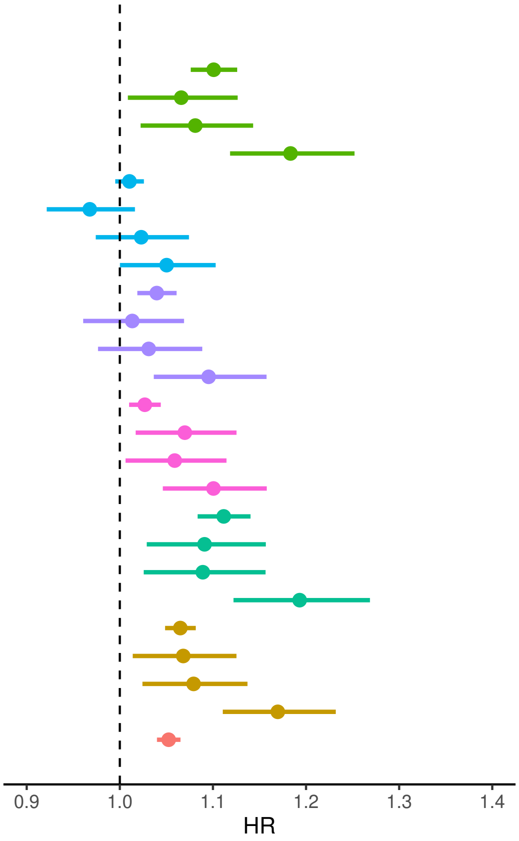

Figure 10: Effect of air pollution on COPD. From left to right, for each air pollution exposure we show its associated hazard ratio and confidence interval, the forest plot and p-value of the effect estimate (Wald test); 'iqr' and '$n^{th}$q' refer to the continuous (scale by IQR) and discrete exposures ($n^{th}$ quartile), respectively. The models were adjusted for age, sex, ethnicity, educational attainment, income, population density, and IMD.

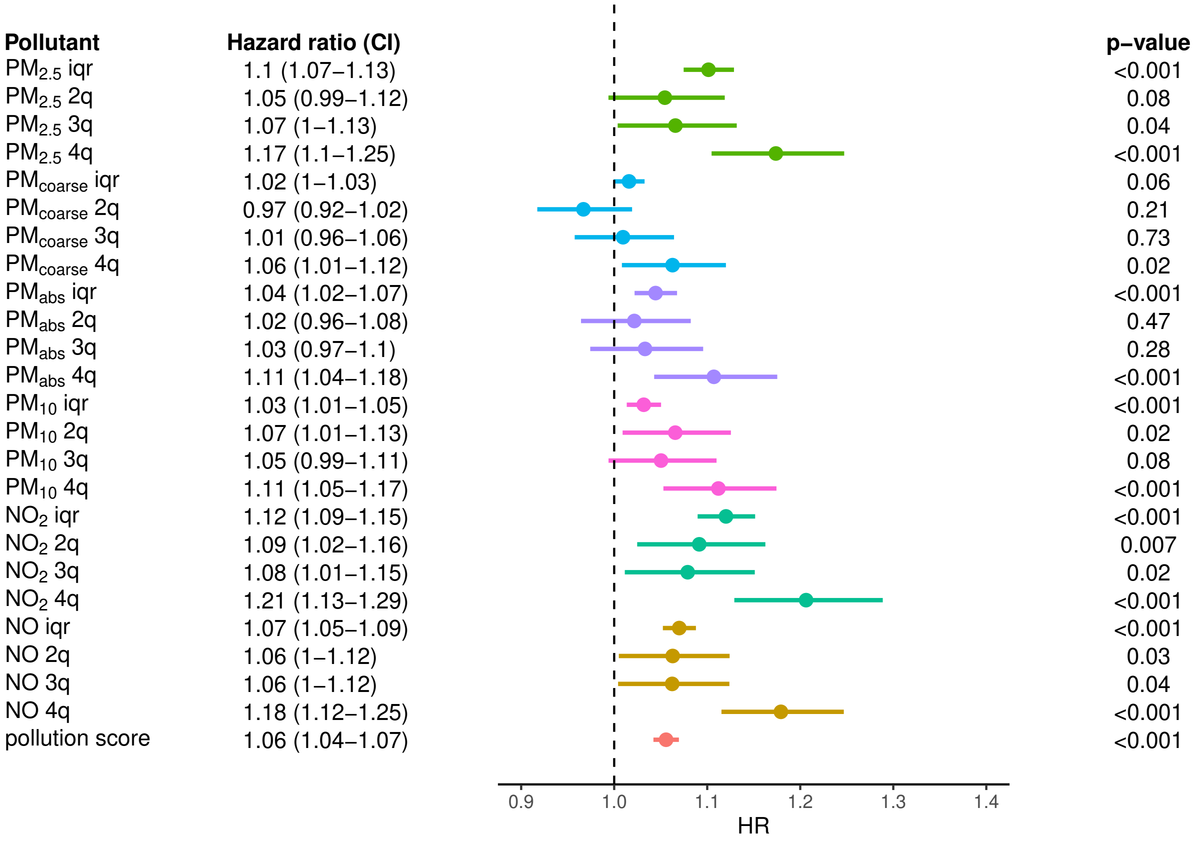

Figure 11: Effect of air pollution on COPD. From left to right, we show the pollution exposure with the associated hazard ratio and confidence interval, the forest plot and p-value of the effect estimate (Wald test); 'iqr' and '$n^{th}$q' refer to the continuous (scale by IQR) and discrete exposures ($n^{th}$ quartile), respectively. The models were adjusted for age, sex, ethnicity, educational attainment, income, population density, and IMD. Additionally, here we excluded participants who lived less than 5 years (instead of 1 year) at baseline address.

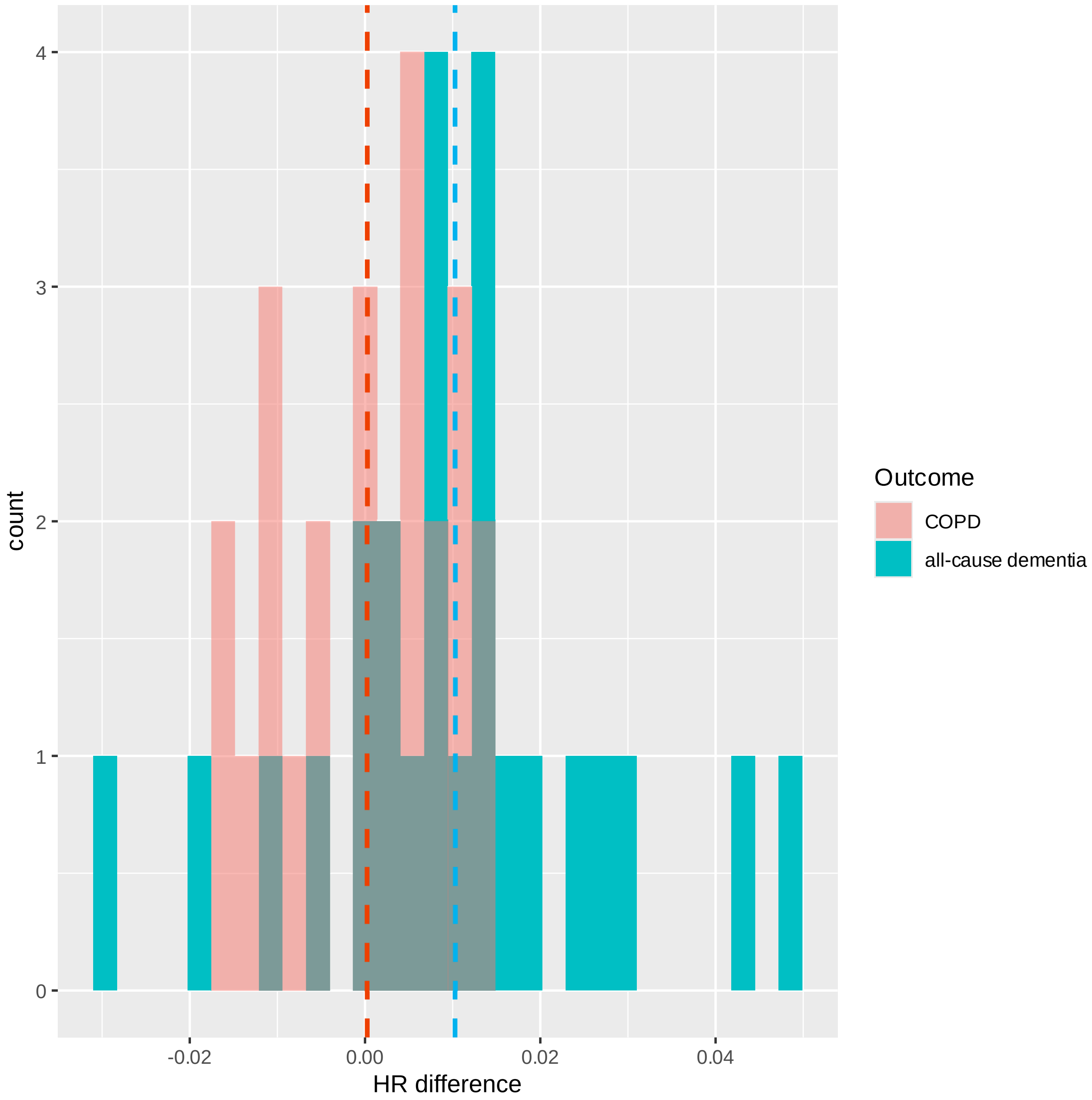

Figure 12: Difference in HR for 5- vs 1-year cutoff. The two histograms show the distribution of differences in hazard ratio (HR) when using 5- vs 1-year cutoff for time lived at baseline address, for all-cause dementia (cyan) and COPD (red) as an outcome. Positive values indicate a higher risk of dementia detected in the analyses using 5-year cutoff. Vertical dashed lines indicate the mean of the distributions. Comparing the two distributions, it appears that there is a larger increase in HR for dementia compared to COPD (Wilcoxon paired test, T=73, p=0.015)

### 7 Negative control analysis

We used ordinal logistic regression to assess whether we could detect an association between air pollution exposure and oily fish intake frequency after adjusting for confounders. Oily fish consumption was coded from higher to lower intake frequency, so that increased odds ratios indicate lower probability of fish intake. Table 4 shows the results for the pollution score and single-pollutant models, with oily fish intake frequency as an ordinal outcome, and exposure encoded continuously, scaled by inter-quartile range; we did not considered air pollution quartiles as an exposure for this analysis. For the pollution score model, the exposure was the first principal component capturing variability in exposure to $PM_{2.5}$, $PM_{abs}$, $NO_2$ and NO.

| | model | OR | L95 | U95 |
|---|---|---|---|---|
| $PM_{2.5}$ | IMD anudjusted | 0.99 | 0.98 | 1.00 |
| | IMD adjusted | 0.97 | 0.97 | 0.98 |
| $PM_{coarse}$ | IMD anudjusted | 0.99 | 0.98 | 0.99 |
| | IMD udjusted | 0.99 | 0.98 | 0.99 |
| $PM_{abs}$ | IMD anudjusted | 0.97 | 0.96 | 0.98 |
| | IMD adjusted | 0.95 | 0.95 | 0.96 |
| $PM_{10}$ | IMD anudjusted | 0.99 | 0.99 | 1.00 |
| | IMD adjusted | 0.99 | 0.98 | 1.00 |
| $NO_2$ | IMD anudjusted | 0.98 | 0.97 | 0.99 |
| | IMD adjusted | 0.96 | 0.95 | 0.97 |
| NO | IMD anudjusted | 0.99 | 0.99 | 1.00 |
| | IMD adjusted | 0.99 | 0.98 | 0.99 |
| pollution score | IMD anudjusted | 0.99 | 0.99 | 0.99 |
| | IMD adjusted | 0.98 | 0.98 | 0.98 |

Table 4: Effect of air pollution of oily fish consumption. Odds ratios lower than one indicate a decrease in probability of consuming less fish, or otherwise an increase in probability of consuming more fish. The models were adjusted for age, sex, ethnicity, educational attainment, income, and population density; additionally, the models could be adjusted for index of multiple deprivation (IMD). L95 and U95 correspond to the lower and upper limit of the 95% confidence interval.

### 8 Control for noise pollution

Figure 13 displays the result from Cox regressions using pollution score and single-pollutant models that included noise pollution as an additional covariate.

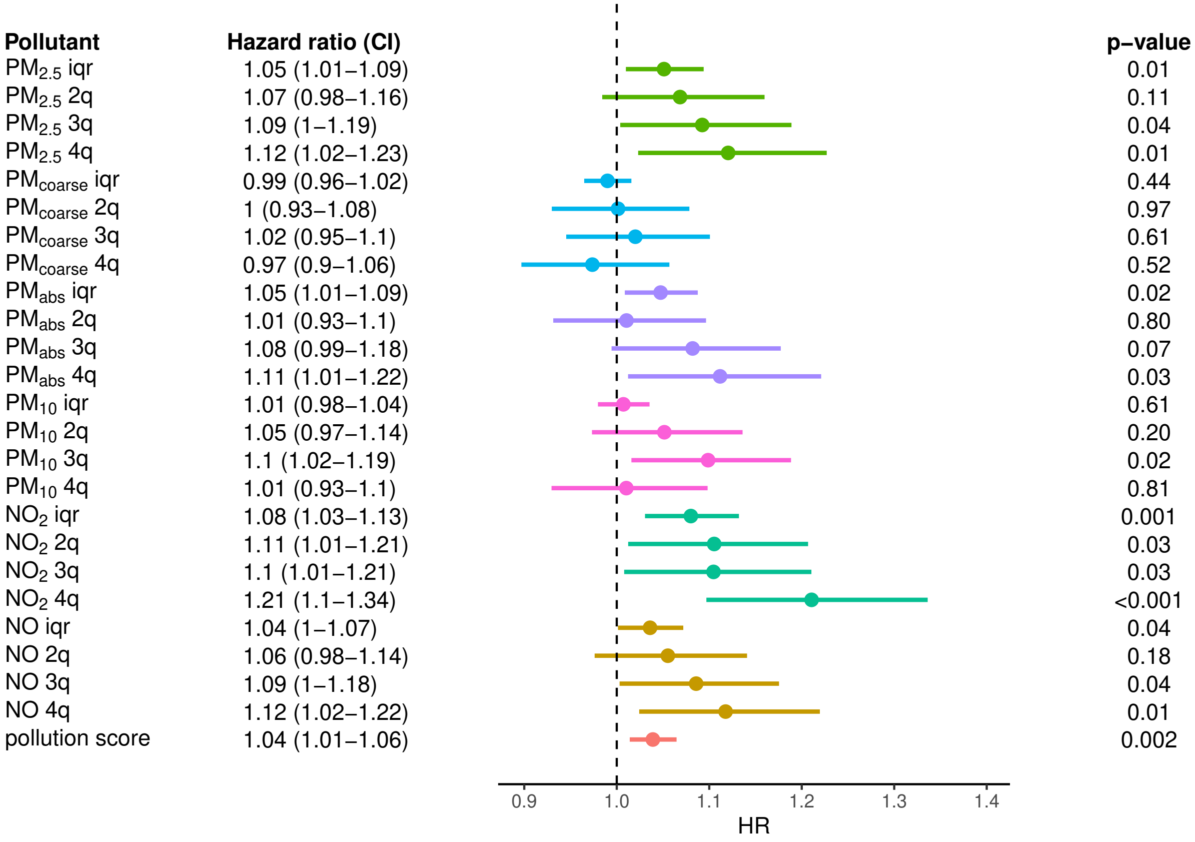

Figure 13: Effect of air pollutants of risk of all-cause dementia when controlling for noise pollution. From left to right, for each air pollution exposure we show its associated hazard ratio and confidence interval, the forest plot and p-value of the effect estimate (Wald test). The models were adjusted for age, sex, ethnicity, educational attainment, income, population density, IMD and noise pollution. Additionally, here we excluded participants who lived less than 5 years (instead of 1 year) at baseline address.

#### 9 Effect modification by deprivation level

We assessed whether the effect of air pollution on dementia is modified by deprivation, both on the multiplicative and additive scale. In the latter case, we used the relative excess risk due to interaction (RERI), which gives the direction of the additive modification. Tables 5-18 report the estimated effect modification on the multiplicative and additive scale, together with the hazard ratio for each combination of exposure and deprivation: in particular, HR10 is the HR for participants with air pollution exposure of 1 iqr above the median and assigned the $1^{st}$ quartile of IMD, HR01 is the HR for participants assigned the $4^{th}$ quartile of IMD and with air pollution exposure equal to the median, and HR11 is the HR for participants both with air pollution exposure of 1 IQR above the medial and assigned the $4^{th}$ quartile of IMD. The interaction on the multiplicative scale is given by $\exp^{\beta_{11}}$=HR11/(HR01*HR10), where $\beta_{11}$ is the interaction coefficient in the linear model of the Cox regression; the RERI is given by HR11-HR10-HR01-1. We ran the analysis both when excluding participants who lived less that 1 year and when excluding participants who less than 5 years at baseline address.

| | Measures | Estimates | L95 | U95 | p-value |
|---|---|---|---|---|---|
| 1 | HR00 | 1.000 | NA | NA | NA |
| 2 | HR01 | 1.264 | 1.167 | 1.370 | 0.000 |
| 3 | HR10 | 1.046 | 0.960 | 1.140 | 0.305 |
| 4 | HR11 | 1.304 | 1.202 | 1.415 | 0.000 |
| 7 | Multiplicative scale | 0.986 | 0.892 | 1.091 | 0.786 |
| 8 | RERI | -0.006 | -0.119 | 0.107 | 0.542 |

Table 5: Effect modification for $PM_{2.5}$. L95 and U95 correspond to the lower and upper limit of the 95% confidence interval. The model was adjusted for adjusted for age, sex, ethnicity, educational attainment, income, population density, and IMD. Hazard ratio (HR), relative excess risk due to interaction (RERI), not a number (NA).

| | Measures | Estimates | L95 | U95 | p-value |
|---|---|---|---|---|---|
| 1 | HR00 | 1.000 | NA | NA | NA |
| 2 | HR01 | 1.249 | 1.146 | 1.360 | 0.000 |
| 3 | HR10 | 1.066 | 0.973 | 1.168 | 0.171 |
| 4 | HR11 | 1.313 | 1.205 | 1.431 | 0.000 |
| 7 | Multiplicative scale | 0.986 | 0.886 | 1.098 | 0.801 |
| 8 | RERI | -0.002 | -0.124 | 0.120 | 0.511 |

Table 6: Effect modification for $PM_{2.5}$. L95 and U95 correspond to the lower and upper limit of the 95% confidence interval. The model was adjusted for adjusted for age, sex, ethnicity, educational attainment, income, population density, and IMD. Additionally, here we excluded participants who lived less than 5 years (instead of 1 year) at baseline address. Hazard ratio (HR), relative excess risk due to interaction (RERI), not a number (NA).

| | Measures | Estimates | L95 | U95 | p-value |
|---|---|---|---|---|---|
| 1 | HR00 | 1.000 | NA | NA | NA |
| 2 | HR01 | 1.286 | 1.188 | 1.393 | 0.000 |
| 3 | HR10 | 1.003 | 0.922 | 1.091 | 0.941 |
| 4 | HR11 | 1.298 | 1.199 | 1.405 | 0.000 |
| 7 | Multiplicative scale | 1.006 | 0.913 | 1.108 | 0.907 |
| 8 | RERI | 0.008 | -0.097 | 0.113 | 0.438 |

Table 7: Effect modification for $PM_{abs}$. L95 and U95 correspond to the lower and upper limit of the 95% confidence interval. The model was adjusted for adjusted for age, sex, ethnicity, educational attainment, income, population density, and IMD. Hazard ratio (HR), relative excess risk due to interaction (RERI), not a number (NA).

| | Measures | Estimates | L95 | U95 | p-value |
|---|---|---|---|---|---|
| 1 | HR00 | 1.000 | NA | NA | NA |
| 2 | HR01 | 1.265 | 1.162 | 1.378 | 0.000 |
| 3 | HR10 | 1.031 | 0.944 | 1.126 | 0.496 |
| 4 | HR11 | 1.305 | 1.200 | 1.419 | 0.000 |
| 7 | Multiplicative scale | 1.000 | 0.903 | 1.107 | 0.999 |
| 8 | RERI | 0.008 | -0.105 | 0.121 | 0.443 |

Table 8: Effect modification for $PM_{abs}$. L95 and U95 correspond to the lower and upper limit of the 95% confidence interval. The model was adjusted for adjusted for age, sex, ethnicity, educational attainment, income, population density, and IMD. Additionally, here we excluded participants who lived less than 5 years (instead of 1 year) at baseline address. Hazard ratio (HR), relative excess risk due to interaction (RERI), not a number (NA).

| | Measures | Estimates | L95 | U95 | p-value |
|---|---|---|---|---|---|
| 1 | HR00 | 1.000 | NA | NA | NA |
| 2 | HR01 | 1.307 | 1.209 | 1.412 | 0.000 |
| 3 | HR10 | 0.984 | 0.937 | 1.032 | 0.504 |
| 4 | HR11 | 1.265 | 1.169 | 1.368 | 0.000 |
| 7 | Multiplicative scale | 0.984 | 0.922 | 1.051 | 0.631 |
| 8 | RERI | -0.025 | -0.100 | 0.049 | 0.749 |

Table 9: Effect modification for $PM_{coarse}$. L95 and U95 correspond to the lower and upper limit of the 95% confidence interval. The model was adjusted for adjusted for age, sex, ethnicity, educational attainment, income, population density, and IMD. Hazard ratio (HR), relative excess risk due to interaction (RERI), not a number (NA).

| | Measures | Estimates | L95 | U95 | p-value |
|---|---|---|---|---|---|
| 1 | HR00 | 1.000 | NA | NA | NA |
| 2 | HR01 | 1.304 | 1.200 | 1.416 | 0.000 |
| 3 | HR10 | 0.993 | 0.944 | 1.045 | 0.783 |
| 4 | HR11 | 1.273 | 1.171 | 1.384 | 0.000 |
| 7 | Multiplicative scale | 0.983 | 0.918 | 1.054 | 0.635 |
| 8 | RERI | -0.024 | -0.102 | 0.055 | 0.721 |

Table 10: Effect modification for $PM_{coarse}$. L95 and U95 correspond to the lower and upper limit of the 95% confidence interval. The model was adjusted for adjusted for age, sex, ethnicity, educational attainment, income, population density, and IMD. Additionally, here we excluded participants who lived less than 5 years (instead of 1 year) at baseline address. Hazard ratio (HR), relative excess risk due to interaction (RERI), not a number (NA).

| | Measures | Estimates | L95 | U95 | p-value |
|---|---|---|---|---|---|
| 1 | HR00 | 1.000 | NA | NA | NA |
| 2 | HR01 | 1.297 | 1.203 | 1.399 | 0.000 |
| 3 | HR10 | 1.012 | 0.960 | 1.065 | 0.665 |
| 4 | HR11 | 1.279 | 1.179 | 1.388 | 0.000 |
| 7 | Multiplicative scale | 0.975 | 0.910 | 1.045 | 0.469 |
| 8 | RERI | -0.030 | -0.109 | 0.049 | 0.770 |

Table 11: Effect modification for $PM_{10}$. L95 and U95 correspond to the lower and upper limit of the 95% confidence interval. The model was adjusted for adjusted for age, sex, ethnicity, educational attainment, income, population density, and IMD. Hazard ratio (HR), relative excess risk due to interaction (RERI), not a number (NA).

| | Measures | Estimates | L95 | U95 | p-value |
|---|---|---|---|---|---|
| 1 | HR00 | 1.000 | NA | NA | NA |
| 2 | HR01 | 1.292 | 1.192 | 1.401 | 0.000 |
| 3 | HR10 | 1.028 | 0.973 | 1.086 | 0.321 |
| 4 | HR11 | 1.286 | 1.180 | 1.402 | 0.000 |
| 7 | Multiplicative scale | 0.968 | 0.899 | 1.042 | 0.383 |
| 8 | RERI | -0.035 | -0.119 | 0.050 | 0.788 |

Table 12: Effect modification for $PM_{10}$. L95 and U95 correspond to the lower and upper limit of the 95% confidence interval. The model was adjusted for adjusted for age, sex, ethnicity, educational attainment, income, population density, and IMD. Additionally, here we excluded participants who lived less than 5 years (instead of 1 year) at baseline address. Hazard ratio (HR), relative excess risk due to interaction (RERI), not a number (NA).

| | Measures | Estimates | L95 | U95 | p-value |
|---|---|---|---|---|---|
| 1 | HR00 | 1.000 | NA | NA | NA |
| 2 | HR01 | 1.264 | 1.163 | 1.373 | 0.000 |
| 3 | HR10 | 1.034 | 0.939 | 1.139 | 0.496 |
| 4 | HR11 | 1.315 | 1.207 | 1.432 | 0.000 |
| 7 | Multiplicative scale | 1.006 | 0.899 | 1.126 | 0.915 |
| 8 | RERI | 0.017 | -0.109 | 0.143 | 0.396 |

Table 13: Effect modification for $NO_2$. L95 and U95 correspond to the lower and upper limit of the 95% confidence interval. The model was adjusted for adjusted for age, sex, ethnicity, educational attainment, income, population density, and IMD. Hazard ratio (HR), relative excess risk due to interaction (RERI), not a number (NA).

| | Measures | Estimates | L95 | U95 | p-value |
|---|---|---|---|---|---|
| 1 | HR00 | 1.000 | NA | NA | NA |
| 2 | HR01 | 1.243 | 1.138 | 1.358 | 0.000 |
| 3 | HR10 | 1.058 | 0.955 | 1.172 | 0.284 |
| 4 | HR11 | 1.327 | 1.213 | 1.452 | 0.000 |
| 7 | Multiplicative scale | 1.009 | 0.896 | 1.137 | 0.880 |
| 8 | RERI | 0.026 | -0.110 | 0.163 | 0.354 |

Table 14: Effect modification for $NO_2$. L95 and U95 correspond to the lower and upper limit of the 95% confidence interval. The model was adjusted for adjusted for age, sex, ethnicity, educational attainment, income, population density, and IMD. Additionally, here we excluded participants who lived less than 5 years (instead of 1 year) at baseline address. Hazard ratio (HR), relative excess risk due to interaction (RERI), not a number (NA).

| | Measures | Estimates | L95 | U95 | p-value |
|---|---|---|---|---|---|
| 1 | HR00 | 1.000 | NA | NA | NA |
| 2 | HR01 | 1.275 | 1.180 | 1.378 | 0.000 |
| 3 | HR10 | 1.017 | 0.947 | 1.093 | 0.635 |
| 4 | HR11 | 1.304 | 1.207 | 1.408 | 0.000 |
| 7 | Multiplicative scale | 1.005 | 0.926 | 1.090 | 0.906 |
| 8 | RERI | 0.011 | -0.078 | 0.100 | 0.402 |

Table 15: Effect modification for NO. L95 and U95 correspond to the lower and upper limit of the 95% confidence interval. The model was adjusted for adjusted for age, sex, ethnicity, educational attainment, income, population density, and IMD. Hazard ratio (HR), relative excess risk due to interaction (RERI), not a number (NA).

| | Measures | Estimates | L95 | U95 | p-value |
|---|---|---|---|---|---|
| 1 | HR00 | 1.000 | NA | NA | NA |
| 2 | HR01 | 1.268 | 1.168 | 1.377 | 0.000 |
| 3 | HR10 | 1.048 | 0.972 | 1.129 | 0.223 |
| 4 | HR11 | 1.301 | 1.199 | 1.412 | 0.000 |
| 7 | Multiplicative scale | 0.980 | 0.899 | 1.067 | 0.637 |
| 8 | RERI | -0.014 | -0.110 | 0.081 | 0.616 |

Table 16: Effect modification for NO. L95 and U95 correspond to the lower and upper limit of the 95% confidence interval. The model was adjusted for adjusted for age, sex, ethnicity, educational attainment, income, population density, and IMD. Additionally, here we excluded participants who lived less than 5 years (instead of 1 year) at baseline address. Hazard ratio (HR), relative excess risk due to interaction (RERI), not a number (NA).

| | Measures | Estimates | L95 | U95 | p-value |
|---|---|---|---|---|---|
| 1 | HR00 | 1.000 | NA | NA | NA |
| 2 | HR01 | 1.266 | 1.167 | 1.373 | 0.000 |
| 3 | HR10 | 1.017 | 0.968 | 1.069 | 0.497 |
| 4 | HR11 | 1.287 | 1.193 | 1.390 | 0.000 |
| 7 | Multiplicative scale | 0.999 | 0.944 | 1.058 | 0.985 |
| 8 | RERI | 0.004 | -0.058 | 0.066 | 0.451 |

Table 17: Effect modification for pollution score. L95 and U95 correspond to the lower and upper limit of the 95% confidence interval. The model was adjusted for adjusted for age, sex, ethnicity, educational attainment, income, population density, and IMD. Hazard ratio (HR), relative excess risk due to interaction (RERI), not a number (NA).

| | Measures | Estimates | L95 | U95 | p-value |
|---|---|---|---|---|---|
| 1 | HR00 | 1.000 | NA | NA | NA |
| 2 | HR01 | 1.245 | 1.142 | 1.358 | 0.000 |
| 3 | HR10 | 1.036 | 0.982 | 1.092 | 0.195 |
| 4 | HR11 | 1.279 | 1.180 | 1.388 | 0.000 |
| 7 | Multiplicative scale | 0.992 | 0.934 | 1.054 | 0.800 |
| 8 | RERI | -0.001 | -0.068 | 0.065 | 0.516 |

Table 18: Effect modification for pollution score. L95 and U95 correspond to the lower and upper limit of the 95% confidence interval. The model was adjusted for adjusted for age, sex, ethnicity, educational attainment, income, population density, and IMD. Additionally, here we excluded participants who lived less than 5 years (instead of 1 year) at baseline address. Hazard ratio (HR), relative excess risk due to interaction (RERI), not a number (NA).

### 10 Proportionality assumption check

We checked the proportionality assumption for Cox regression using Schoenfeld residuals. In figure 14 we plot the linear fit to the residuals obtained from the main model, using all cause-dementia as an outcome and with all pollutants, scaled by interquartile range, as exposure. Comparable results were obtained from single pollutant models, as exemplified in Figure 15 for $PM_{2.5}$. We found that proportionality assumptions were violated for age, sex, income, $PM_{10}$, $PM_{2.5}$, $PM_a bsorbance$ and $NO_2$ ($p < 0.05$), but that results were likely due to the high sample size: in fact, no clear violation was visible when plotting the residuals (Figure 16), especially for air pollutants. Moreover, the results did not change when regressing all cause dementia on exposure to pollution score when stratifying the effect of age, sex, and income by time (Table 19): this indicates that our effect estimates for air pollution were likely constant over time-in-study, thus respecting the proportionality assumption. Here we split time in two using a cut-point at 12 years, based on the visual inspection of the order 3 polynomial fits, shown in Figure 17, as suggested by [9].

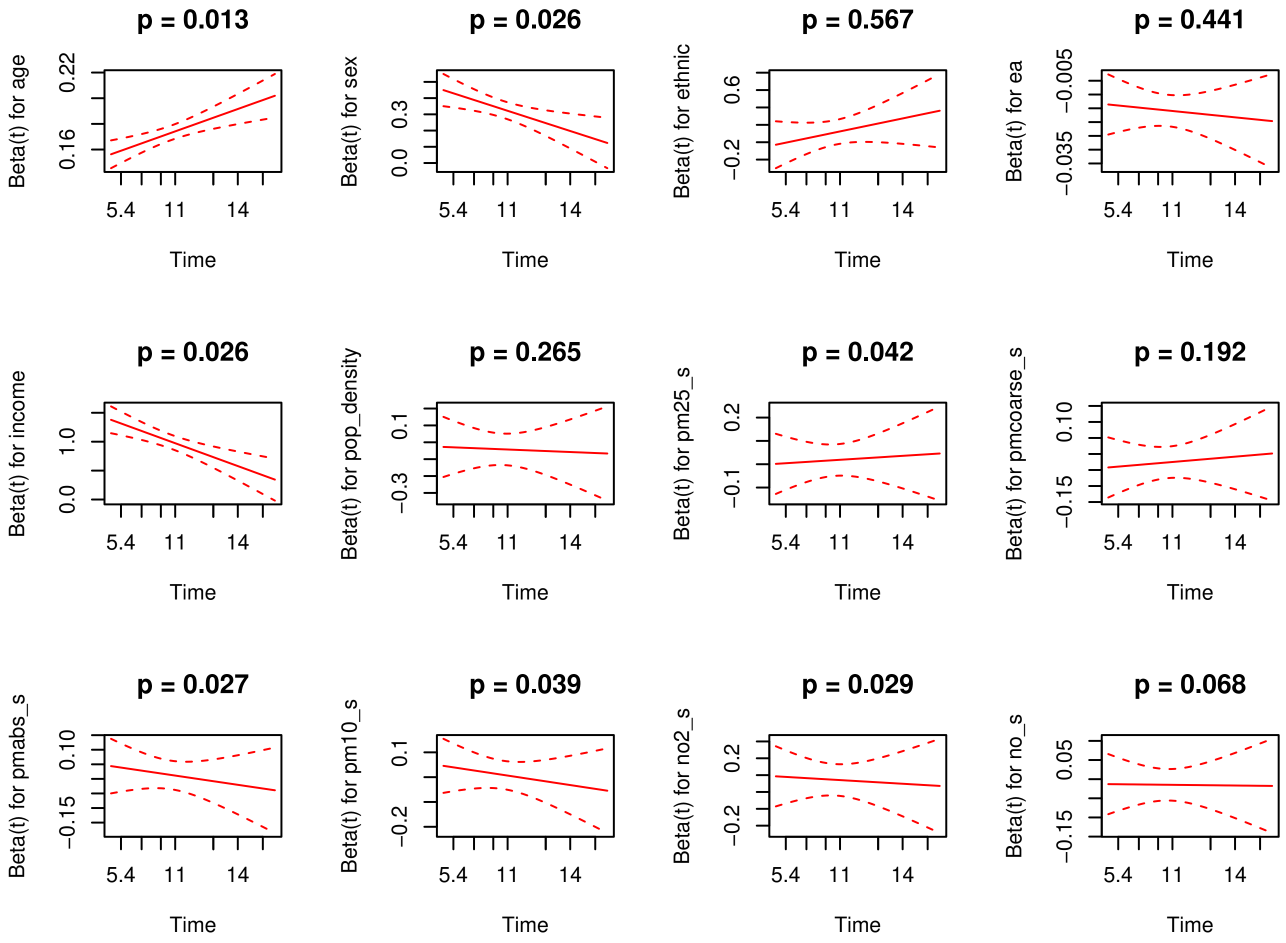

Figure 14: Linear fit, and associated p-values, to Schoenfeld residuals. The model was adjusted for age, sex, ethnicity, educational attainment, income, and population density, the outcome was all-cause dementia and the exposure included all air pollutants.

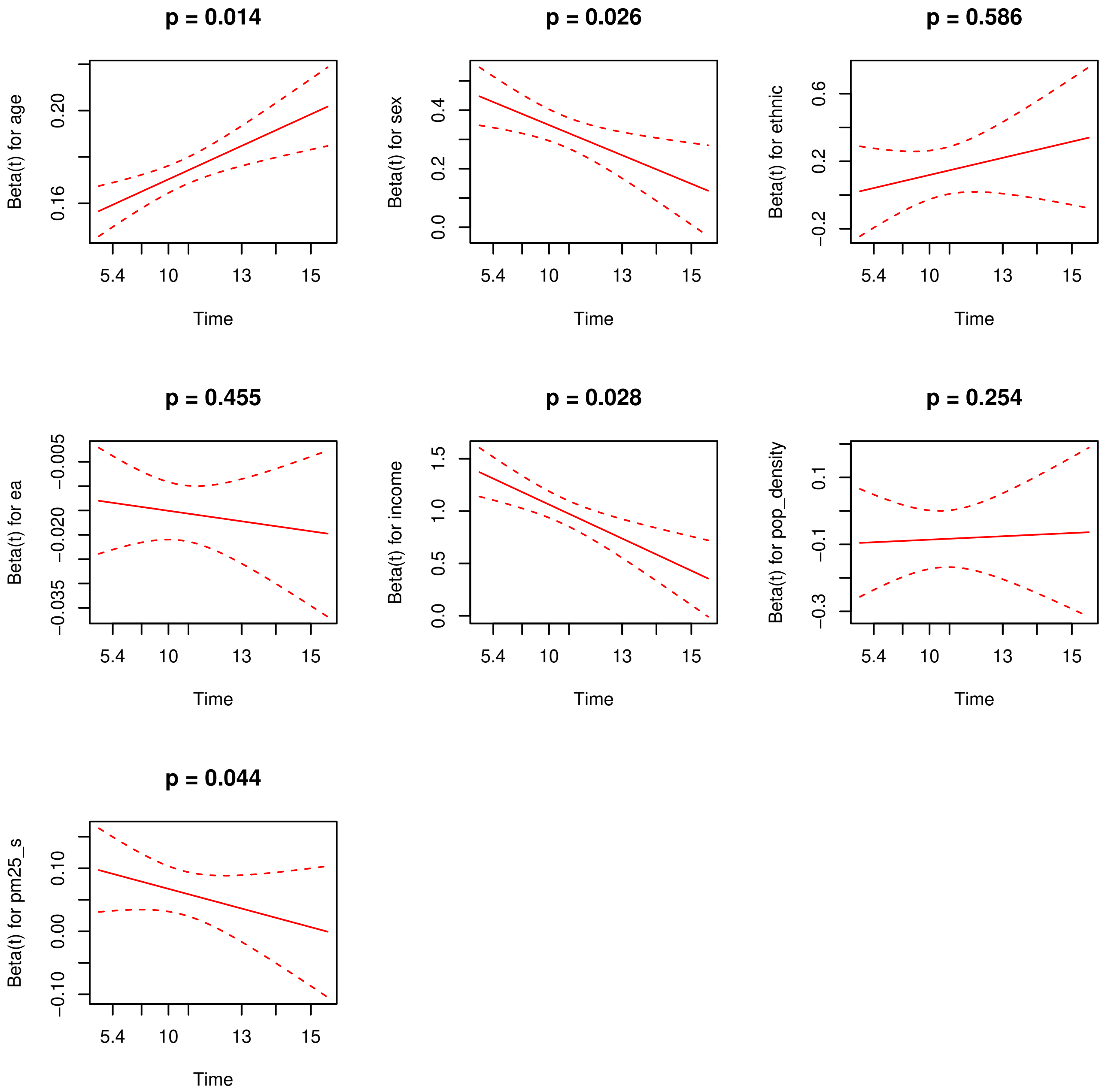

Figure 15: Linear fit, and associated p-values, to Schoenfeld residuals. The model is the same as in Figure14, but with only $PM_{2.5}$ as exposure.

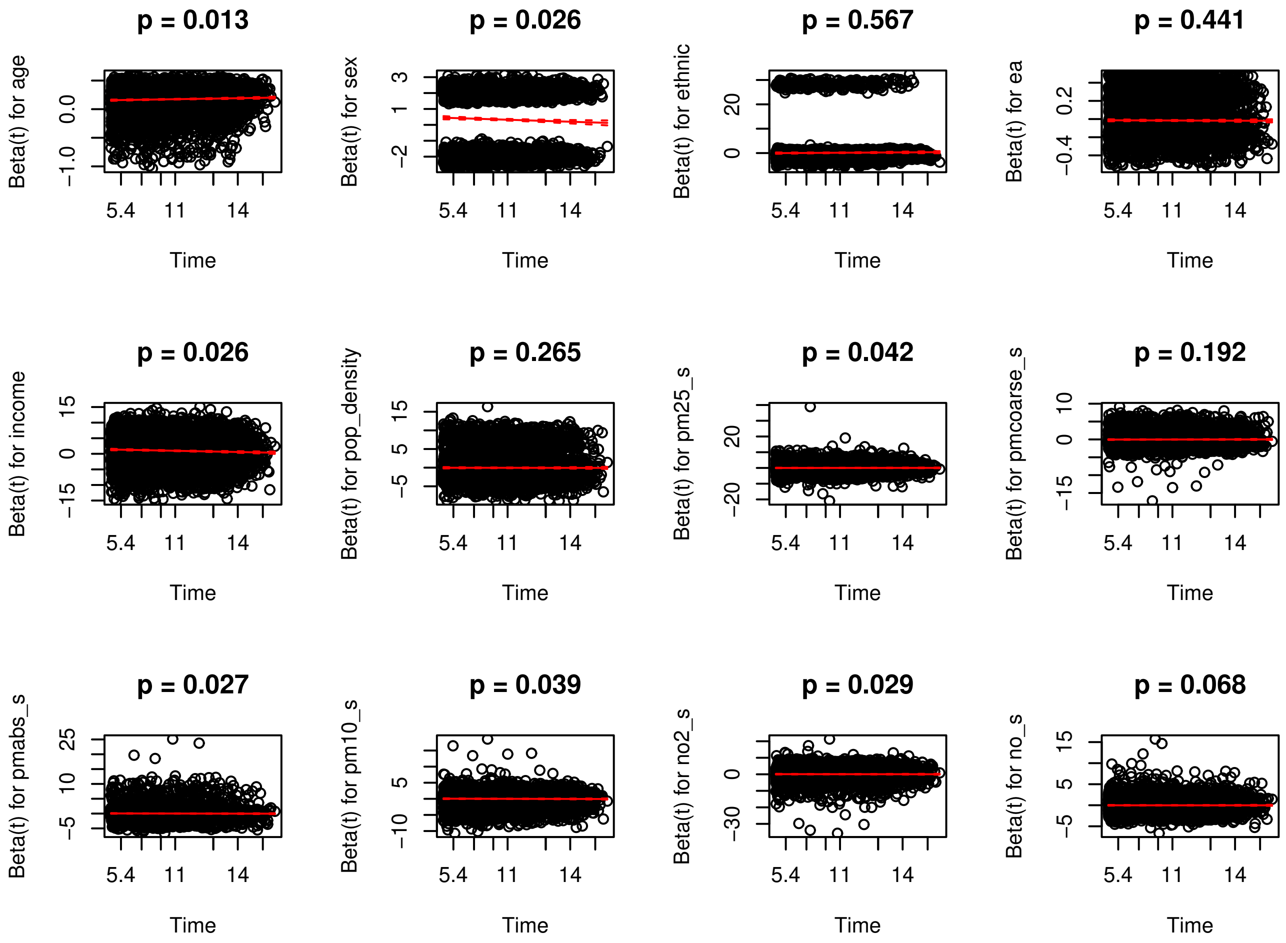

Figure 16: Plot of Schoenfeld residuals together with the linear fit. The model is the same as in Figure14

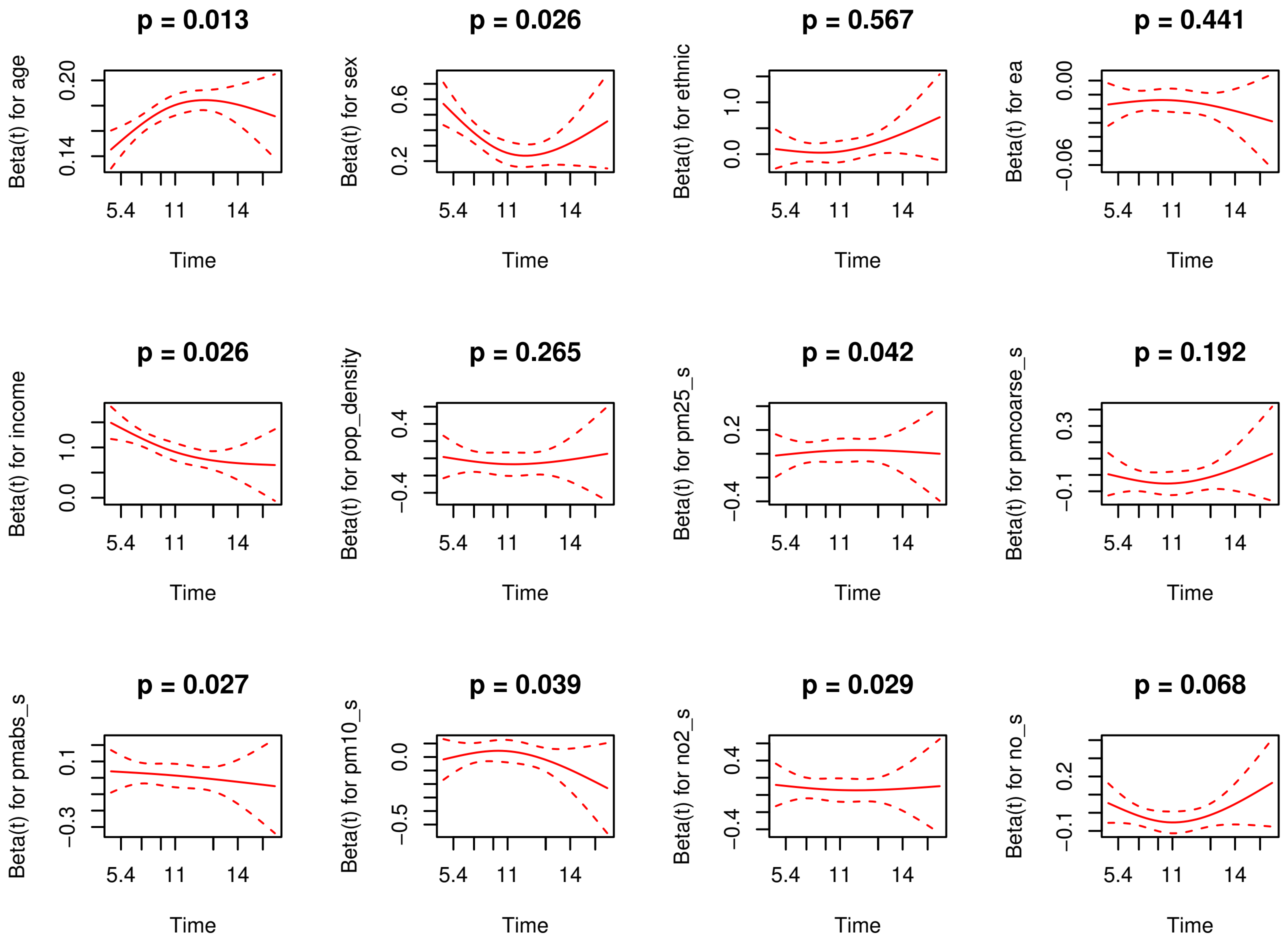

Figure 17: Degree 3 polynomial fit to Schoenfeld residuals. The model is the same as in Figure14. The fits highlight a consistent change in slope approximately around 12 years.

| model | HR | L95 | U95 | p-value |
|---|---|---|---|---|
| pollution score | 1.0348 | 1.0162 | 1.0537 | $<$0.001 |
| pollscore-age | 1.0348 | 1.0162 | 1.0537 | $<$0.001 |
| pollution score-sex | 1.0348 | 1.0162 | 1.0537 | $<$0.001 |
| pollution score-income | 1.0349 | 1.0164 | 1.0538 | $<$0.001 |

Table 19: Comparison of hazard ratio for air pollution score for different models. Here we used the main model (adjusted for age, sex, ethnicity, education, income, and population density) with pollution score as an exposure as reference. The other models included respectively an interaction between time period and age, sex, and income. The hazard ratio for pollution score for the different models is invariant, suggesting that the effect estimates in our analysis were constant over time-in-study.

### 11 Air pollutants characteristics

| Pollutant | Min. | 1st Q | Median | Mean | 3rd Q | Max. |
|---|---|---|---|---|---|---|
| $pm_{2.5}$ | 8.170 | 9.280 | 9.920 | 9.973 | 10.550 | 21.250 |
| $pm_{coarse}$ | 5.570 | 5.840 | 6.100 | 6.411 | 6.620 | 12.820 |
| $pm_{abs}$ | 0.830 | .990 | 1.130 | 1.182 | 1.300 | 4.570 |
| $pm_{10}$ | 11.78 | 15.23 | 16.02 | 16.21 | 16.98 | 30.65 |
| $NO_2$ | 12.93 | 21.31 | 26.07 | 26.56 | 31.17 | 108.49 |
| NO | 0 | 11.61 | 15.94 | 17.23 | 20.53 | 160.06 |

Table 20: Distribution of air pollution data. Units are in $\mu g/m^3$. Q is quartile

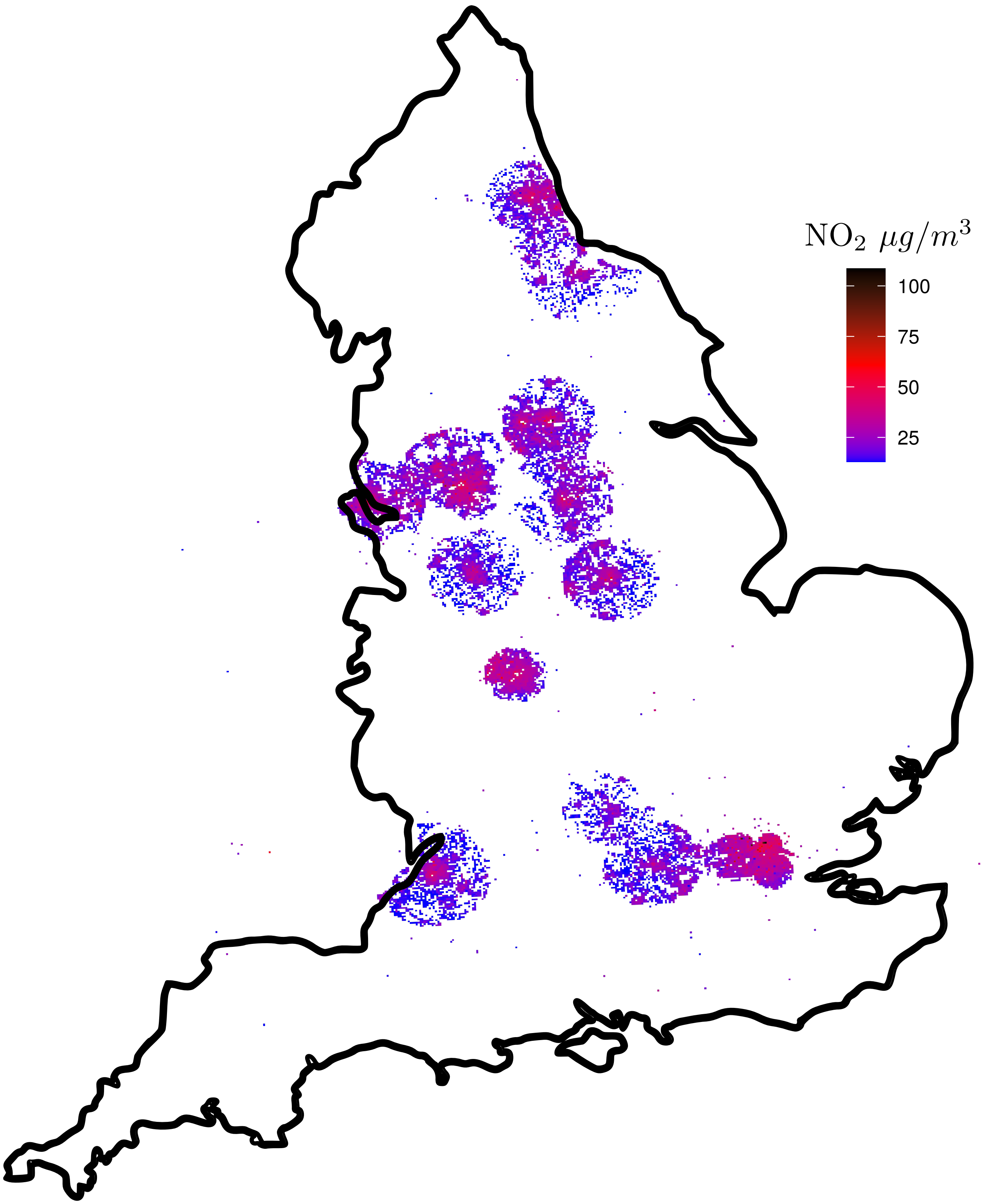

Figure 18: Spatial distribution of $NO_2$ exposure across participants considered in this study

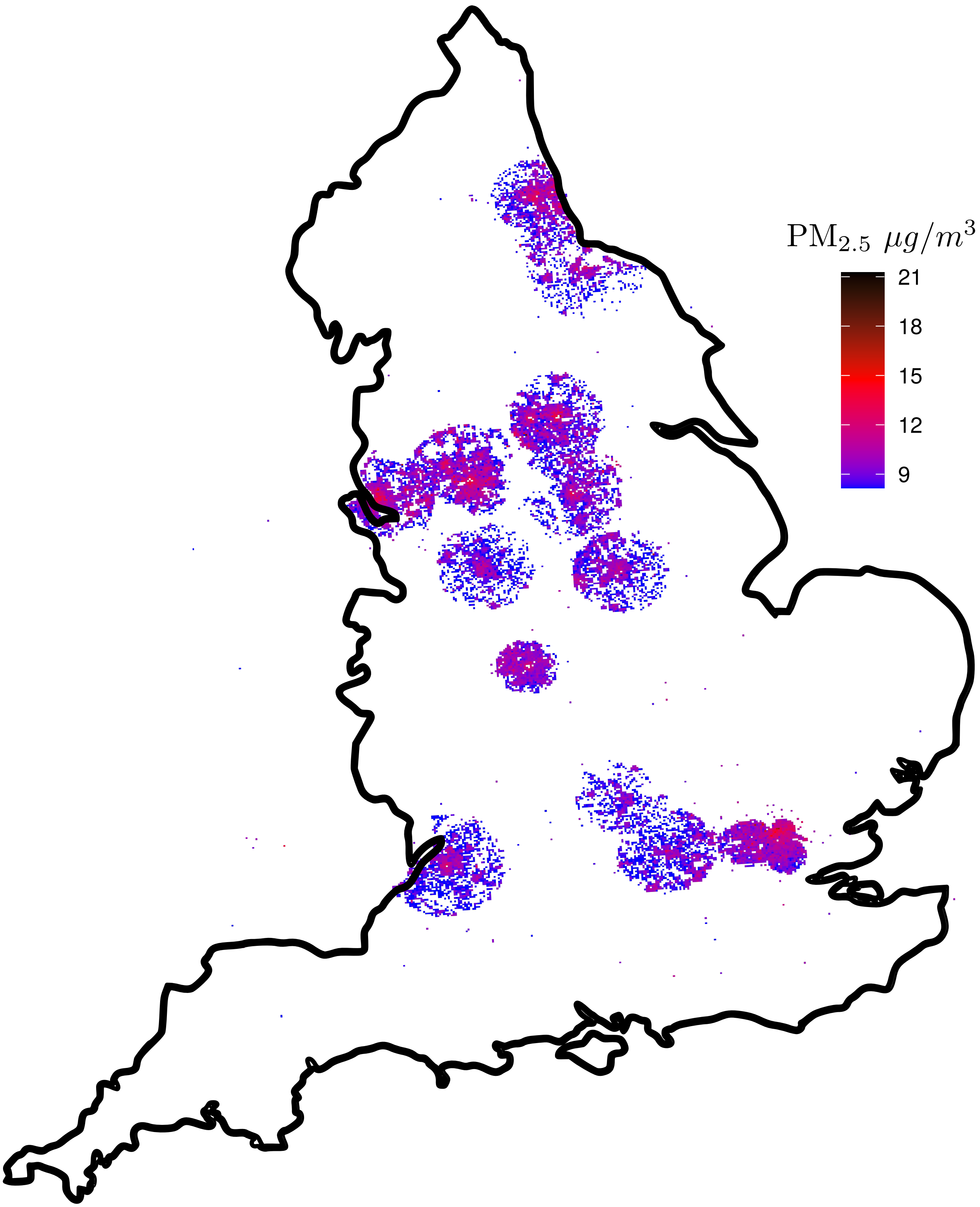

Figure 19: Spatial distribution of $PM_{2.5}$ exposure across participants considered in this study

| | $PM_{2.5}$ | $PM_{coarse}$ | $PM_{abs}$ | $PM_{10}$ | $NO_2$ | NO |
|---|---|---|---|---|---|---|
| $PM_{2.5}$ | 1.00 | 0.22 | 0.60 | 0.54 | 0.86 | 0.73 |
| $PM_{coarse}$ | 0.22 | 1.00 | 0.42 | 0.81 | 0.20 | 0.24 |
| $PM_{abs}$ | 0.60 | 0.42 | 1.00 | 0.56 | 0.75 | 0.49 |
| $PM_{10}$ | 0.54 | 0.81 | 0.56 | 1.00 | 0.51 | 0.46 |
| $NO_2$ | 0.86 | 0.20 | 0.75 | 0.51 | 1.00 | 0.74 |
| NO | 0.73 | 0.24 | 0.49 | 0.46 | 0.74 | 1.00 |

Table 21: Pearson correlation between air pollutants
